# Genetics of Cardiac Aging Implicate Organ-Specific Variation

**DOI:** 10.1101/2024.08.02.24310874

**Authors:** James Brundage, Joshua P. Barrios, Geoffrey H. Tison, James P. Pirruccello

## Abstract

Heart structure and function change with age, and the notion that the heart may age faster for some individuals than for others has driven interest in estimating cardiac age acceleration. However, current approaches have limited feature richness (heart measurements; radiomics) or capture extraneous data and therefore lack cardiac specificity (deep learning [DL] on unmasked chest MRI). These technical limitations have been a barrier to efforts to understand genetic contributions to age acceleration. We hypothesized that a video-based DL model provided with heart-masked MRI data would capture a rich yet cardiac-specific representation of cardiac aging.

In 61,691 UK Biobank participants, we excluded noncardiac pixels from cardiac MRI and trained a video-based DL model to predict age from one cardiac cycle in the 4-chamber view. We then computed cardiac age acceleration as the bias-corrected prediction of heart age minus the calendar age. Predicted heart age explained 71.1% of variance in calendar age, with a mean absolute error of 3.3 years. Cardiac age acceleration was linked to unfavorable cardiac geometry and systolic and diastolic dysfunction. We also observed links between cardiac age acceleration and diet, decreased physical activity, increased alcohol and tobacco use, and altered levels of 239 serum proteins, as well as adverse brain MRI characteristics.

We found cardiac age acceleration to be heritable (h2g 26.6%); a genome-wide association study identified 8 loci related to linked to cardiomyopathy (near *TTN, TNS1, LSM3, PALLD, DSP, PLEC, ANKRD1* and *MYO18B*) and an additional 16 loci (near *MECOM, NPR3, KLHL3, HDGFL1, CDKN1A, ELN, SLC25A37, PI15, AP3M1, HMGA2, ADPRHL1, PGAP3, WNT9B, UHRF1 and DOK5*). Of the discovered loci, 21 were not previously associated with cardiac age acceleration. Mendelian randomization revealed that lower genetically mediated levels of 6 circulating proteins (MSRA most strongly), as well as greater levels of 5 proteins (LXN most strongly) were associated with cardiac age acceleration, as were greater blood pressure and Lp(a). A polygenic score for cardiac age acceleration predicted earlier onset of arrhythmia, heart failure, myocardial infarction, and mortality.

These findings provide a thematic understanding of cardiac age acceleration and suggest that heart- and vascular-specific factors are key to cardiac age acceleration, predominating over a more global aging program.

## Introduction

The heart undergoes a decline in function with age including atrial enlargement and concentric left ventricular (LV) remodeling ^1,2^. At a cellular level, cardiac aging has been characterized by disabled macroautophagy, loss of proteostasis, genomic instability and epigenetic alterations contributing to mitochondrial dysfunction, dysregulated neurohormonal signaling, and inflammation ^3^. While the number of senescent cells increases with age, this appears to be variable across tissues ^4^. Further, the aging process does not appear to occur at equal rates over chronological time for all people. For example, DNA methylation clocks have estimated calendar age in cells from multiple tissue types, with “ticks” of the clock moving faster during periods of development and stress ^5,6^. There are some whose hearts have features consistent with an age older than their chronological age, which has been termed cardiac age acceleration^7^. Better characterization of cardiac age acceleration may help to identify individuals at risk of early heart disease and to identify druggable pathways to decelerate or reverse cardiac aging.

Cardiac age acceleration has been studied in adults using data ranging from proteomics to electrocardiography (ECG) and magnetic resonance imaging (MRI). Many sources demonstrate associations with lifestyle factors such as tobacco use, alcohol consumption, adiposity; and to downstream cardiovascular disease risk^7–10^.

Several studies have linked cardiac age acceleration to common genetic variants, though genetic characterizations of cardiac age acceleration are influenced by the data type and modeling approach used to estimate age. For example, Se-Hwee Oh, *et al*, found evidence linking plasma proteomic markers to cardiac age acceleration ^11^. Libiseller-Egger, *et al*, estimated cardiac age acceleration from ECG and linked this measure to seven genetic loci at genome-wide significance, including those near *TTN* and *SCN5A*/*SCN10A* ^10^. Shah, *et al*, used standard cardiovascular measurements derived from MRI to estimate cardiac age acceleration in the UK Biobank ^9^, additionally linking genetic loci near *ELN*, *PI15*, and *PLCE1* to cardiac age acceleration. These loci had little overlap with those from ECG-estimated cardiac age acceleration, which might be expected if the component of cardiac age that can be embedded in the ECG differs from that in the cardiac MRI. Raisi-Estabragh, *et al*, expanded this analysis to incorporate radiomics features from MRI ^8,12^. Goallec, *et al*, generalized this to an end-to-end deep learning model incorporating both ECG and MRI videos. Despite explaining more variance in age and achieving a high heritability of 37.9% for age acceleration (**Supplementary Table 1**), this end-to-end deep learning model—trained on videos containing the heart and surrounding tissues such as adipose, lungs, skeletal muscle, and bones—identified fewer loci for cardiac age acceleration, including one locus near *TTN* and three intergenic loci^7^.

In the present work, we sought to derive a model of age acceleration in which the model is blinded to non-cardiac structures, hypothesizing that such an approach would enhance the cardiovascular specificity of genetic contributions to cardiac age acceleration. In particular, we hypothesized that a two-stage deep learning approach would combine the strengths of cardiac-specific data with the flexible learning capabilities of an end-to-end deep learning model: first, by training a model to mask out non-cardiac tissues from MRI CINEs, and second, by estimating cardiac age acceleration from those masked images.

## Methods

### Study population

The study was conducted within the UK Biobank, a cohort of 502,638 participants recruited from 2006 to 2010, aged 40-70 at recruitment. An imaging substudy was ongoing at the time of this analysis, and at the time of model training, 70,805 participants had MRI data available. Analyses were considered exempt by the UCSF Institutional Review Board (IRB), #22-37715. UK Biobank analyses were conducted under application #41664. Baseline characteristics of the cohort used for training and validation of the model can be found in **Supplemental Table 2**.

### MRI Preprocessing

The long-axis 4-chamber cardiac MRI images in the UK Biobank were represented as 208×208 pixel images, with 50 images over the course of one cardiac cycle per imaging substudy participant. The pixel values were stored with up to 16 bits of range (0-65,535). Prior to analysis, these pixel values were reformatted into the 0-1 range, and any image that was not originally 208×208 pixels was zero-padded to that dimension. While some participants underwent imaging at two separate imaging visits, only data from the first imaging visit was analyzed for each participant.

### MRI Segmentation

198 four-chamber long-axis cardiac MRI stillframe images were manually segmented by a cardiologist (JPP) and used to train a deep learning model to perform semantic segmentation. A deep learning image segmentation model was constructed using a U-Net architecture ^13^. The PyTorch version 2.0.1 built-in implementation of ConvNeXt-Small was used as the encoder for the model, with weights that had been pre-trained on ImageNet1K (obtained from PyTorch’s torchvision package) ^14,15^. The encoder was modified to accept 1-channel input, instead of its default 3-channel input, by averaging the kernel weights in the input layer. At each of the four downsampling layers of the ConvNeXt encoder, a skip connection was created to the corresponding upsampling layer of the decoder. Each layer of the decoder used a pixelshuffle step followed by convolution, batchnorm, and the GELU non-linearity ^16^. The skip connections from the corresponding downsampling layer of the encoder were incorporated using an attention-block to incorporate the skip-connection data from the encoder with the output of the prior layer of the decoder. All non-pretrained layers were initialized with PyTorch’s *kaiming_normal* function. The loss function for training was the linear combination of focal loss and Dice loss at a 20:1 ratio described in *Segment Anything* ^17^. The *AdamW* optimizer with weight decay 0.0001 was used with the PyTorch *OneCycleLR* cyclic learning rate scheduler, with learning rate ranging from 1e-5 to a maximum or 2e-3 during training^18^.

### Cardiac Age Acceleration

We next jointly used both the raw imaging data and the segmentation output to measure accelerated heart aging. A deep learning model was trained to regress age from cardiac MRI CINEs masked to include only cardiac tissue: specifically, all pixels in each image that were not within the atria or ventricles were set to a pixel intensity value of 0. The trained model was then used to generate predicted age for each MRI included in the dataset.

The calendar age was then subtracted from predicted age, resulting in a value termed the *cardiac age delta*. A common problem faced in age regression problems, including cardiac age and brain age, is that trained models tend to overestimate age for participants below the mean age, and underestimate age for participants above the mean age, a concept termed regression dilution bias. Here, we adjust for regression dilution bias, similar to Raisi-Estabragh et al., and we refer to this adjusted value as *cardiac age acceleration* (**Supplemental Figure 1A-C**). To correct the cardiac age acceleration values, a linear regression model was used to estimate the cardiac age delta in the training set via equation 1:

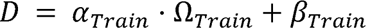

where *D* is the cardiac age delta, *α_Train_* is the slope, Ω*_Train_* represents calendar age and *β_Train_* represents the intercept. From the formula, it is evident that age prediction can occur in the absence of information about calendar age, but age acceleration requires calendar age. The alpha and beta parameters were then used to generate a predicted cardiac age delta from each participant in the test set, which was subtracted from the predicted heart age to correct for regression dilution bias via equation 2:

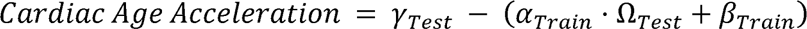

where *γ_Test_* is predicted heart age, *α*_Train_ and *β_Train_* are slope and intercept from the training set equation 1 respectively, and Ω*_Test_* is calendar age of the test set participant. This process was performed for each fold cross validation.

### Dataset Splitting/Cross Validation

The dataset was split into multiple training, testing and validation groups for different experiments, detailed below.

The goal of model cross validation was to generate predictions for every participant in the dataset, without the model being contaminated by data in the training set. To accomplish this, the dataset was split into a training/testing set of 61,691 participants, a validation set of 3,195 participants, and a smaller holdout set of 674 participants. 5,245 participants had MRIs become available after the initial training process, and were added to the analysis, but were not included in the original dataset split: these were included as a second holdout set and contributed to downstream analyses but not to model evaluation.

The training/testing set was subdivided into 10 groups, G0-G9. During model training, each one of the groups was excluded from the training data in turn. This resulted in an approximate 90:10 ratio of training to testing MRIs during model training, building a total of 10 models. For all 10 models, the shared validation set was used to monitor model progress and select the best-fit weights for each of the 10 models. Following training, inference was performed using each model on the group that was not included in its training set. For example, for the G0 fold, there were 55,571 MRIs in the training set (consisting of groups 1-9). During training, that model was validated against the shared validation set, and then applied to the 6,190 MRIs in the G0 set. This process was repeated for each of the 10 models.

By using this approach, each participant received a prediction from a model that did not use its data for training—an approach designed to minimize overfitting. To ensure these models performed consistently, each model was also tested on the holdout set that was not used in any of the training, testing, or validation described above (**Supplemental Table 3**). For downstream analyses, CINEs included in the holdout and validation sets used predictions from the G0 fold model.

### Cardiac Age Model Training

The age prediction model was trained using the PyTorch 1.13.1 library in python^15^. Baseline augmentations were carried out in PyTorch, and included zero-padding the MRI CINEs to shape 208×208 pixels with 50 frames. MRIs from the training set were further augmented using Kornia 0.7.0 augmentations, by rotating the heart in a range of ±180 degrees, with the translation set between 0.0 and 0.5 and altering the CINE with a gamma between 0.5 and 2.0 ^19^. A ResNet18 based R2 Plus 1D convolutional neural network (CNN), with weights initialized from training on the Kinetics dataset was used, as detailed by Tran et al. ^20^. Architecture details and validation can be found in Tran et al. In brief, the architecture took a 4-dimensional tensor as input in the form 1 (single color channel) x *Number of Frames* x *Height* x *Width*, followed by 16 (2+1)D convolutional blocks, a global average pooling layer and a fully connected layer. Each (2+1)D block was comprised of a 2D spatial convolutional layer with a 3×3 kernel size, batch normalization, rectified linear unit (ReLU) followed by a 1D temporal convolutional layer, another batch normalization and ReLU. The architecture with Kinetics weights are publicly available via the PyTorch library.

Models were trained with a mean squared error (MSE) loss function, ADAM optimizer and a one cycle learning rate scheduler ^21^. The initial learning rate was set at 0.001 and was selected by a learning rate finder process ^18^. Weight decay was set at 0.0001. Automatic mixed precision was used to accelerate model training. Gradient clipping was also used via PyTorch’s *clip_grad_norm_* to improve regularization and prevent exploding gradients with a max norm of 1 and norm type 2.

Besides the cardiac specific model mentioned above, a model was also trained without masking, allowing it to learn from cardiac and non-cardiac structures. The unmasked model was trained only with a single fold of the training and testing set. All other training methods including architecture, hyperparameters and loss were all the same as the cardiac specific model.

### Model Evaluation

The models were evaluated using mean absolute error (MAE), the mean of the absolute value of the difference between the true age and predicted age. They were also evaluated by Pearson correlation, a measure of linear correlation between the predicted age and the calendar age. Finally, they were also evaluated by R^2^, which is the square of the Pearson correlation coefficient for a one-parameter model with intercept.

### Alignment Filtering

To assess MRI quality, an augmentation-based approach was used to identify MRIs that likely had poor alignment of all four heart chambers, or poor segmentation of the heart. To estimate MRI quality, an MRI and its associated segmentation mask underwent a rotational transformation. The rotated MRI was then used to generate a new segmentation mask for each frame of each CINE. A DICE score was calculated between the original rotated segmentation mask and the new segmentation mask generated from the rotated MRI were compared for all four heart chamber segmentations for each frame ^22^. The mean of those DICE scores was calculated for each CINE. The resultant mean DICE scores for the four heart chambers were then used in principle component analysis (PCA). All MRIs with a component 1 or component 2 greater than one standard deviation from the mean were visualized and given a quality score of pass or fail. By this standard 10,054 MRIs were manually visualized and rated. As the value of each principal component decreased, a higher percentage of MRIs received a passing quality score. 2,872 participants received a failing MRI alignment quality score and were excluded from GWAS analysis. An overview of this process is displayed in **Supplemental Figure 2**.

### Arrhythmia Filtering

To avoid detecting age acceleration due to simply identifying abnormal cardiac motion from arrhythmia, participants whose 12-lead ECG (obtained during the imaging visit on the same day as the MRI) reported an automated read of atrial fibrillation or flutter were excluded (1,412 excluded).

### Adjusting for MRI Derived Phenotypes

To determine the extent to which standard cardiac measurements (e.g., left ventricular ejection fraction) could account for cardiac age acceleration, linear regression was performed in participants with both imaging-derived phenotypes (from UK Biobank category 157) and cardiac age acceleration measurements. These measurements included 85 total cardiac measurements, including volumes from all four chambers, wall thickness and strain at up to 16 points throughout the left ventricle. For a complete list of phenotypes included, see **Supplemental Table 4** The variance in cardiac age acceleration explained by the classical measurements was assessed.

### Continuous PheWAS

To explore associations between continuous phenotypes collected in the UK Biobank and cardiac age acceleration, a linear model was fit between continuous phenotypes and cardiac age acceleration including MRI serial number, sex and age at MRI acquisition as covariates. 11,399 phenotypes were tested (a phenome-wide association study [PheWAS]). Categories of phenotypes tested included serum measurements, measurements extracted from imaging, measurements of diet, lifestyle and activity. They also included levels of 1,463 serum proteins collected at enrollment and determined by the Olink proteomics assay ^23^. Results were adjusted for multiple comparisons using the false discovery rate (FDR) method, with the FDR threshold set at 0.05. For investigation of serum biomarkers, we use the Bonferroni threshold to select a subset of biomarkers for further exploration.

### Disease PheWAS

Associations between prevalent disease and disease incident to the MRI were also investigated. For prevalent disease, a linear model was fit for each disease recorded in the UK Biobank and cardiac age acceleration including MRI serial number, age at MRI and sex as covariates. For incident disease, a Cox model was fit using the same covariates. Results were adjusted for multiple comparisons using the FDR method, with the FDR threshold set at 0.05. UKBiobank fields and ICD codes used to create disease definitions are included in **Bulk Data Table 7**.

### Genome Wide Association Study

Common genetic variants that contribute to the cardiac age acceleration phenotype were discovered using REGENIE version 3.2.7 with imputed variants provided by the UK Biobank. Quality control based on genetic information was performed per the **Supplemental Methods**. In brief, REGENIE runs in two steps; the fitting of a whole genome model from a subset of markers and then the testing of a larger set of markers conditional on the predictions of the step 1 model using a leave one chromosome out (LOCO) approach to minimize proximal contamination. GWAS covariates included age, sex, the MRI serial number, the genotyping array and the first ten principal components of ancestry.

### MAGMA

Tissue data were taken from GTEx v8 ^24,25^. scRNA-seq data for left ventricular cardiomyocytes were taken from Chaffin, et al after reprocessing by PlaqView ^26^. GTEx v8 RNA sequencing gene reads were combined with the GTEx sample attributes metadata and then processed in EdgeR ^27^. The edgeR::DGEList function was called to ingest the data. Then the edgeR::calcNormFactors function was called for normalization. Genes with an expression count of less than 100 were then removed. The data were loaded into voom ^28^. Iterating over each tissue as a target (with weight 1), all other tissues were considered non-target and weighted by −1 times the number of samples contributing to that tissue divided by the total number of samples. Tissues with the same prefix (e.g., “Artery”) were excluded from the non-target tissue set for each target that had a prefix. The design matrix compared expression between the non-target tissues (assigned to negative weights) and the target tissue (assigned to weight +1), with model fitting performed by voom::eBayes. A gene set was created for each target tissue by filtering to keep genes with log fold change between target and non-target tissues of greater than two and false discovery rate (FDR)-adjusted P-value < 0.01.

### Olink cis-pQTL identification

Cis-lead SNPs from Sun, et al, ascertained in 35,571 European-ancestry participants from the random baseline cohort (batches 1-6), were previously described^23^. Sample filtering included retaining variants with INFO > 0.7, minor allele count > 50, and excluding samples with sex mismatch, sex chromosome aneuploidy, and excess heterozygosity. GWAS was performed with REGENIE v2.2.1^29^. A genome-wide Bonferroni correction was applied (5E-08 accounting for 1,463 proteins), retaining SNPs with P < 3.4E-11. Cis SNPs were defined as being within 500kb of the gene encoding region of their respective protein. These significant cis pQTLs (from Sun et al Supplementary Table 6) were retained as exposure markers for Mendelian randomization. Olink assay information can be found in **Supplemental Methods**.

### Trait Exposure Identification

For each of the selected traits (**Supplemental Methods)**, a GWAS was performed in UK Biobank participants excluding those who underwent imaging, and those within 3 degrees of freedom of participants with imaging, in REGENIE v2.2.1^29^. The lead SNPs of each GWAS were used as the exposure markers for Mendelian randomization.

### Mendelian randomization

Mendelian randomization (MR) was performed for each of the Olink proteins and traits listed above using the TwoSampleMR R package v0.5.8^30^. For single-variant traits, the Wald ratio was the only method used. For multi-variant traits, weighted median, inverse variace weighted, Egger, weighted mode and simple mode methods were tested. In addition, Steiger testing was performed to assess whether the variant was more strongly associated with its instrument than the outcome (to reduce the risk of identifying relationships driven by reverse causation or pleiotropy)^31^.

### Polygenic Risk Score

A polygenic score for the heritable component of cardiac age acceleration was constructed using 1.1 million HapMap3 SNPs with *PRScs-auto* using a linkage disequilibrium reference panel that was made publicly available by the *PRScs* authors (https://www.dropbox.com/s/t9opx2ty6ucrpib/ldblk_ukbb_eur.tar.gz) ^32,33^. In *auto* mode, *PRScs* estimates polygenic score weights without requiring cross-validation for hyperparameter tuning in an additional test set.

## Results

### Performance of cardiac age model on segmented heart CINEs

We first trained a deep learning model to identify all cardiac pixels in four-chamber long-axis MRI CINEs, and used that model’s output as a mask to exclude non-cardiac structures when training a second deep learning model to estimate calendar age. The masking ensured that the deep learning model could not use non-cardiac data in the MRI to estimate cardiac age. 70,805 individuals received a cardiac age acceleration measurement (mean 0.0, standard deviation 3.6, **Figure 1**). During the 10-fold cross validation, the models achieved an average MAE of 3.28 and RMSE of 4.17 (**Supplemental Table #5**). Calendar age was associated with predicted age with an average Pearson R of 0.843 and R^2^ of 0.71 (**Supplemental Table #5**, **Supplemental Figure 1A**). These metrics were comparable when each model was tested on the universal holdout set (**Supplemental Table #3**). For more information regarding the impact of adjustment for regression dilution bias, see **Supplemental Results**.

**Figure 1:**
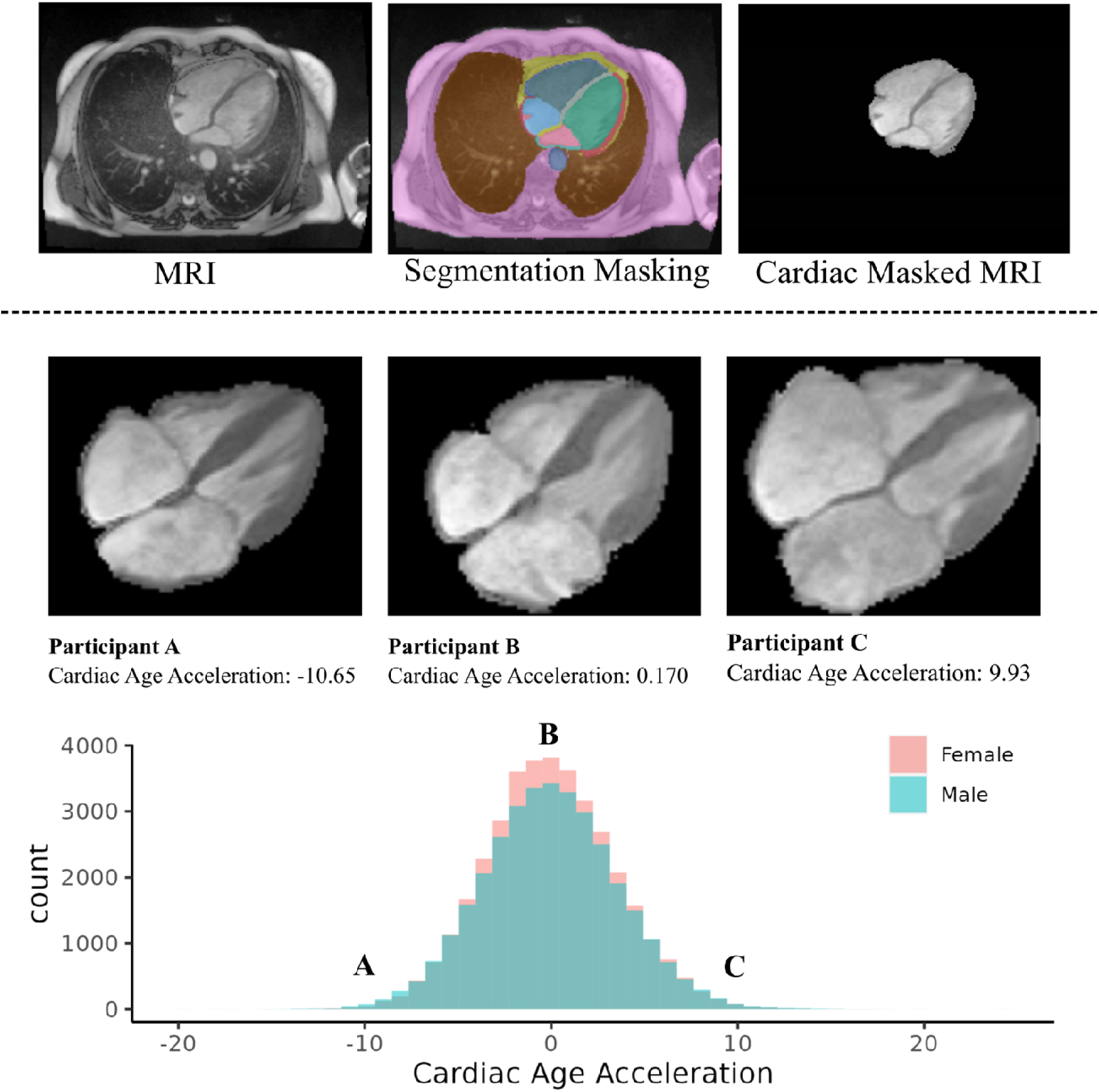
Study overview. *Top:* Representative four chamber full MRI, the same MRI with segmentation applied, and the MRI with non-cardiac structures masked out. *Bottom:* Representative examples from individuals of the same age and sex with cardiac age deceleration (A), no cardiac age acceleration (B) and cardiac age acceleration (C). Heart images reproduced by kind permission of UK Biobank ©.

We sought to understand the degree to which the deep learning-derived cardiac age acceleration measurements could have been accounted for more simply by using common clinical measurements derived from the MRI, such as left ventricular ejection fraction (LVEF) or left ventricular mass. In a subset of up to 30,031 participants with cardiac MRI-derived measurements, a linear model including all of the MRI-derived measurements (see Methods) accounted for 10.4% of the variability in cardiac age acceleration, suggesting that only a small portion of cardiac age acceleration could be accounted for with standard clinical measurements.

### Continuous Trait PheWAS of Cardiac Age Acceleration Summary

We then sought to understand the relationship between cardiac age acceleration and other measurements including anthropometric values, biomarkers, measures of activity and behavior, and imaging-derived phenotypes. Cardiac age acceleration was associated with continuous measurements obtained in the UK Biobank. A total of 66,521 MRIs passed alignment and arrhythmia quality control (**Supplemental Figure #3**). Of the 11,399 continuous phenotypes compared, 2,803 had significant associations with cardiac age acceleration after FDR correction. Like previous literature, cardiac age acceleration was associated with elevated systolic blood pressure (0.146 standard deviation (SD) change per SD increase in cardiac age acceleration, p=4.4E-18) and diastolic blood pressure (0.164 SD, p=1.9E-24) ^7,9^. All other results in the continuous PheWAS will be reported as the SD change in the phenotype per SD change in cardiac age acceleration. The continuous PheWAS results can be found in **Bulk Data Table #1** and are highlighted in the following sections.

### Continuous PheWAS of Cardiac MRI Image Derived Phenotypes

Cardiac age acceleration was associated with structural and functional changes detectable on cardiovascular MRI, including altered systolic and diastolic function and increased chamber sizes (**Figure 2, Supplemental Data Table 1**). In the left ventricle (LV), cardiac age acceleration was associated with increased global radial strain (0.064 SD, p=8.5E-30), global longitudinal strain (0.019 SD, p=4.0E-04), wall thickness (0.100 SD, p=5.2E-52) and ejection fraction (0.02 SD, p=1.0E-3), and decreased global circumferential strain (−0.015 SD, p=6.2E-03), end diastolic volume (−0.080 SD, p=3.2E-33), end systolic volume (−0.060 SD, p=8.8E-21) and stroke volume (−0.064 SD, p=9.2E-26). The same trends were seen in the right ventricle (RV) (**Figure 2, Supplemental Data Table 1**). In the left atrium (LA) it was associated with signs of decreased function, including associations with increased minimum volume (0.03 SD, p=1.3E-08), decreased ejection fraction (−0.05 SD, p=4.9E-21) and stroke volume (−0.06 SD, p=2.0E-29). In the right atrium (RA), it was associated with increased maximum volume (0.02 SD, p=4.7E-3) and stroke volume (0.02 SD, p=1.2E-3). In the ascending aorta it was associated with decreased distensibility (−0.16 SD, p= 5.0E-129) and increased maximum area (0.15 SD, p=3.1E-146). Similar significant changes were seen in the descending aorta (**Figure 2, Supplemental Data Table 1**).

**Figure 2:**
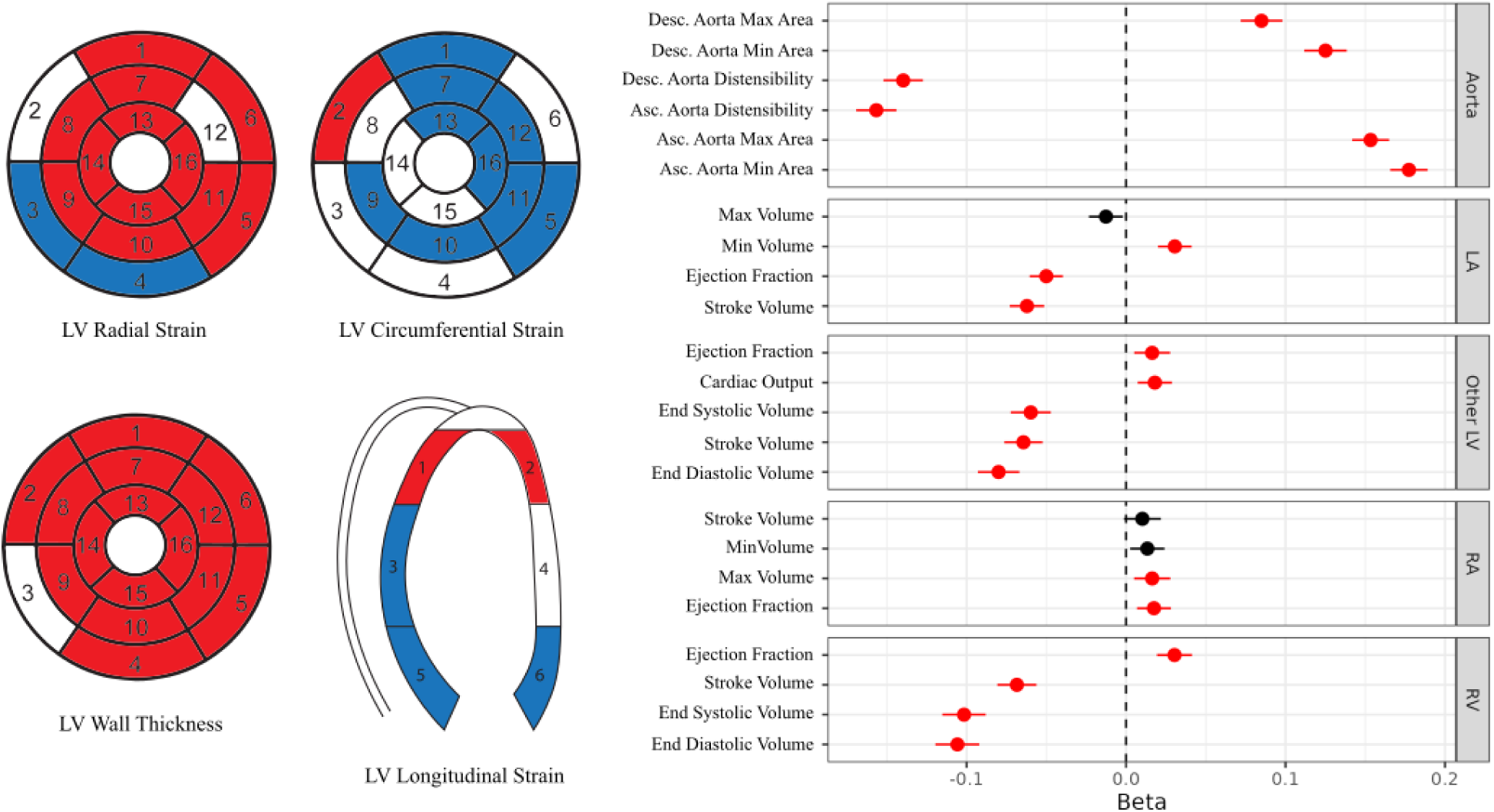
Cardiac structural associations. *Left:* Associations between LV radial strain, circumferential strain, longitudinal strain and wall thickness displayed by AHA segments. Red segments had a significant positive association while blue segments had a significant inverse association. *Right:* A plot of effect sizes between cardiac age acceleration and image derived phenotypes from the cardiac MRI. Red dots reach statistical significance after FDR correction. Beta: standard deviation change in each measurement per standard deviation change in cardiac age acceleration.

**Table 1:** Lead SNPs from the GWAS of cardiac age acceleration.

| SNP | CH | BP | EA | OA | Beta | P value | Nearest Gene |
| --- | --- | --- | --- | --- | --- | --- | --- |
| rs2042995 | 2 | 179558366 | T | C | -0.093 | 8.4E-46 | TTN |
| rs10165160 | 2 | 218694962 | C | G | -0.033 | 1.3E-08 | TNS1 |
| rs7612736 | 3 | 14276238 | G | A | 0.046 | 7.7E-12 | LSM3 |
| rs144856067 | 3 | 168698634 | A | AT | 0.050 | 1.5E-08 | MECOM |
| rs11383131 | 3 | 169177556 | A | AC | -0.036 | 2.3E-10 | MECOM |
| 4:169726307_GA_G | 4 | 169726307 | GA | G | -0.031 | 2.4E-08 | PALLD |
| rs7733331 | 5 | 32828846 | T | C | -0.039 | 6.2E-12 | NPR3 |
| rs61511837 | 5 | 137066469 | C | T | -0.044 | 6.3E-10 | KLHL3 |
| 6:7491551_AT_A | 6 | 7491551 | AT | A | -0.032 | 2.8E-08 | DSP |
| rs140386312 | 6 | 22619557 | G | GT | -0.049 | 1.8E-14 | HDGFL1 |
| rs3176326 | 6 | 36647289 | G | A | 0.056 | 3.6E-16 | CDKN1A |
| rs113395463 | 7 | 73478524 | A | G | 0.077 | 2.3E-18 | ELN |
| rs17089329 | 8 | 23390046 | C | G | -0.044 | 1.8E-10 | SLC25A37 |
| rs993318 | 8 | 75786660 | A | G | 0.043 | 1.8E-14 | PI15 |
| rs11786896 | 8 | 145018354 | C | T | -0.077 | 1.9E-09 | PLEC |
| rs10762576 | 10 | 75896601 | A | C | 0.039 | 2.7E-10 | AP3M1 |
| rs10748555 | 10 | 92682060 | C | A | 0.040 | 4.1E-13 | ANKRD1 |
| rs10878359 | 12 | 66404624 | T | C | -0.032 | 1.4E-08 | HMGA2 |
| rs41306688 | 13 | 114078558 | A | C | -0.110 | 2.5E-13 | ADPRHL1 |
| rs2517955 | 17 | 37843681 | C | T | 0.044 | 2.5E-14 | PGAP3 |
| rs75230966 | 17 | 44967530 | G | A | -0.077 | 1.2E-19 | WNT9B |
| rs2779165 | 19 | 4915447 | G | C | 0.039 | 3.9E-08 | UHRF1 |
| rs2206909 | 20 | 53270624 | C | T | 0.035 | 3.7E-08 | DOK5 |
| rs133889 | 22 | 26160105 | G | C | -0.035 | 1.8E-10 | MYO18B |

SNP dbSNP identifier where available; CH chromosome; BP GRCh37 base position; EA effect allele; OA other allele; Beta REGENIE effect size of the effect allele, in units of the rank-based inverse normal transform which approximates a standard deviation change; P-value two-tailed REGENIE p value.

### Continuous PheWAS of Lifestyle Factors and Activity

Lifestyle factors were significantly associated with cardiac age acceleration. Consumption of substances known to increase risk of heart disease were associated with cardiac age acceleration: acceleration was linked to greater tobacco use (0.09 SD per pack year of smoking, p=2.5E-24) and alcohol consumption (0.08 SD per weekly standard drink, p= 1.2E-42). Similarly, it was associated with increased body mass index (BMI) (0.023 SD, p=1.0E-07), waist circumference (0.046 SD, p=2.5E-23) and multiple measures of fat content on abdominal MRI (**Supplemental Figure 4, Supplemental Data Table 2**).

**Table 2:**
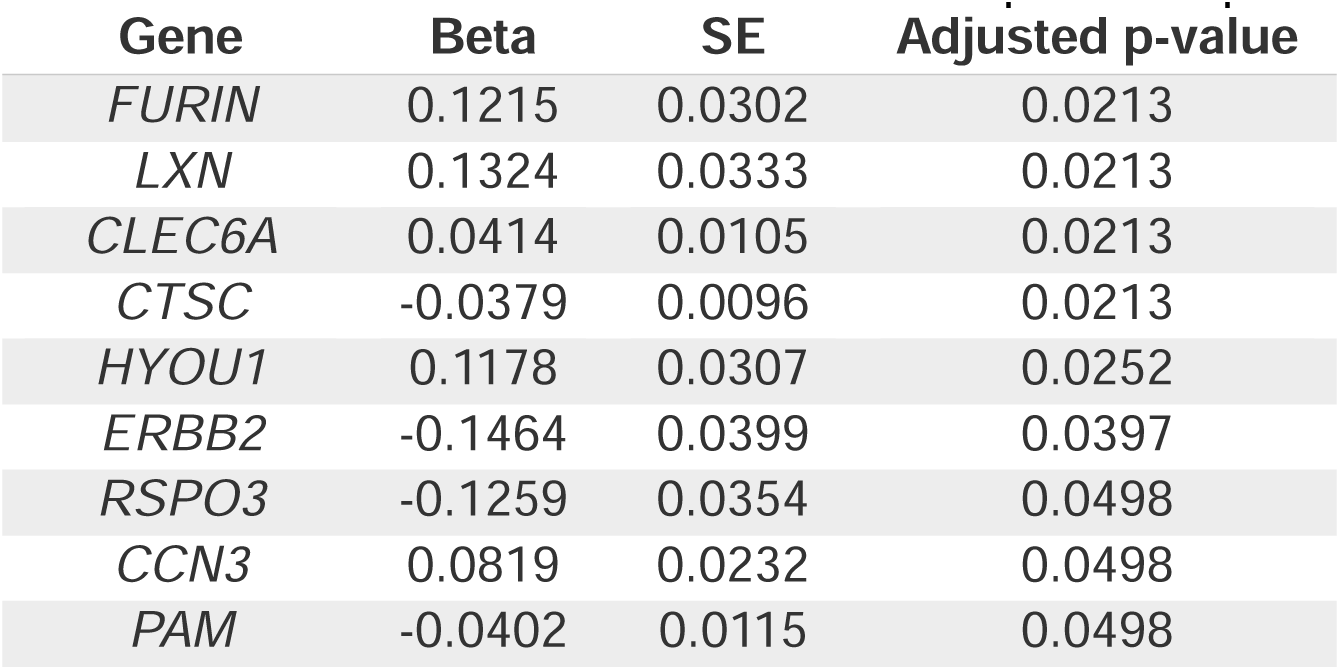
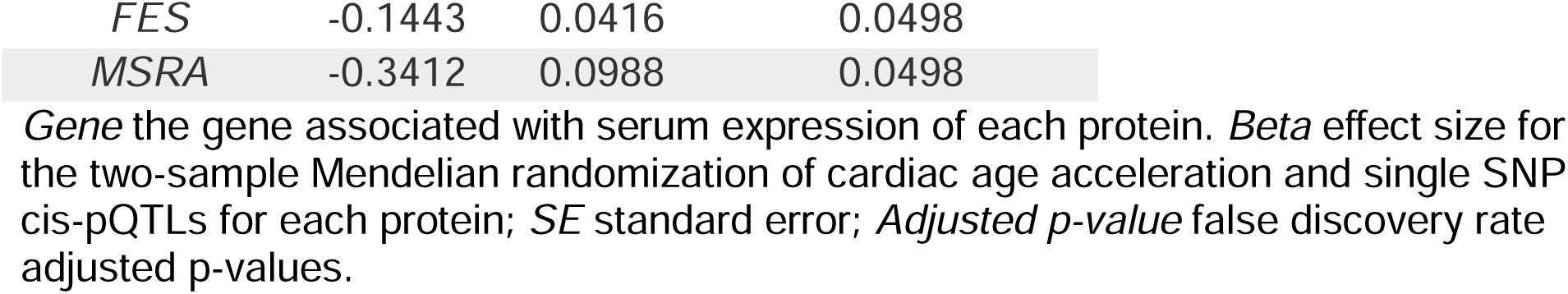
Mendelian randomization of serum protein expression.

| Gene | Beta | SE | Adjusted p-value |
| --- | --- | --- | --- |
| FURIN | 0.1215 | 0.0302 | 0.0213 |
| LXN | 0.1324 | 0.0333 | 0.0213 |
| CLEC6A | 0.0414 | 0.0105 | 0.0213 |
| CTSC | -0.0379 | 0.0096 | 0.0213 |
| HYOU1 | 0.1178 | 0.0307 | 0.0252 |
| ERBB2 | -0.1464 | 0.0399 | 0.0397 |
| RSPO3 | -0.1259 | 0.0354 | 0.0498 |
| CCN3 | 0.0819 | 0.0232 | 0.0498 |
| PAM | -0.0402 | 0.0115 | 0.0498 |
| *FES* | -0.1443 | 0.0416 | 0.0498 |
| *MSRA* | -0.3412 | 0.0988 | 0.0498 |

*Gene* the gene associated with serum expression of each protein. *Beta* effect size for the two-sample Mendelian randomization of cardiac age acceleration and single SNP cis-pQTLs for each protein; *SE* standard error; *Adjusted p-value* false discovery rate adjusted p-values.

Cardiac age acceleration was inversely associated with physical activity measured by wearable accelerometry (**Supplemental Data Table 3**), including moderate to vigorous physical activity on wearable accelerometry (−0.05 SD, p=7.0E-14)^34^. In contrast, cardiac age acceleration was associated with increased “sedentary” activity (0.02, p=6.1E-4) ^34^. In a separate linear model, cardiac age acceleration was lower in a group completing greater than 150 min/week of moderate to vigorous physical activity (MVPA) after adjustment for age and sex (−6.3e-04 SD, p=4.5E-14). This held true even in those whose activity occurred over only 1-2 days out of the week (a “weekend warrior” pattern; −6.3E-4 SD, p=1.7e-13) ^35^. Finally, self-reported increased duration of sleep was associated with cardiac age deceleration (−0.14 SD, p=4.3e-4).

### Continuous PheWAS of Brain MRI and Carotid Ultrasound Image Derived Phenotypes

We examined associations of cardiac age acceleration with changes on brain MRI. Generally, brain MRI derived phenotypes of brain aging were associated with cardiac age acceleration. The T1 MRI showed associations between cardiac age acceleration and decreased volume of grey matter (−9.30E-02 SD, p= 1.9E-44) and increased CSF volume (5.50E-02 SD, p=2.1E-21), which is likely related to grey matter atrophy (**Supplemental Figure 5, Supplemental Data Table 4**). It was associated with decreased volume of white matter, though the association is weaker (−2.10E-02 SD, p=6.6E-05) (**Supplemental Figure 5, Supplemental Data Table 4**). Age acceleration was also associated with increased volume of hyperintensities on T2 imaging (3.80E-02 SD, p= 8.9E-14), with the strongest effect size in the periventricular region (6.80E-02 SD, p=6.4E-35) (**Supplemental Figure 6, Supplemental Data Table 4).** It was associated with decreased cerebral blood flow in all brain regions on arterial spin imaging (**Supplemental Figure 7, Supplemental Data Table 4**). Cardiac age acceleration was also associated with increased quantitative susceptibility mapping (QSM) from susceptibility weighted MRI in the caudate, hippocampus, putamen, pallidum and thalamus but not in the nucleus accumbens or substantia nigra (**Supplemental Figure 8, Supplemental Data Table 4**). It was also associated with carotid artery intimal media thickness on carotid ultrasound (**Supplemental Figure 9, Supplemental Data Table 4**).

We hypothesized that systolic blood pressure may be a shared cardiac and brain aging risk factor that may contribute to the above associations. A sensitivity analysis which repeated the continuous PheWAS but included systolic blood pressure as a covariate was still significant for associations between the brain MRI phenotypes and cardiac age acceleration (**Supplemental Data Table 4**). Inclusion of systolic blood pressure increased the model R^2^ suggesting that if systolic blood pressure mediates associations between cardiac age acceleration and brain imaging phenotypes, it is only partially (**Supplemental Data Table 4**).

### Continuous PheWAS of Circulating Blood Markers and Metabolites

Cardiac age acceleration was also associated with markers of greater bone marrow function, including higher levels of circulating hemoglobin (0.066 SD, p= 1.9E-36), reticulocytes (0.029 SD, p= 9.8E-13), monocytes (0.039 SD, p= 6.3E-22) and neutrophils (0.034 SD, p= 1.3E-17) (**Supplemental Data Table 5, Supplemental Figure 10**).

Cardiac age acceleration was associated with circulating markers of cardiometabolic risk, including higher levels of hemoglobin A1c (0.051 SD, p= 6.5E-35) and glucose (0.046 SD, p= 8.7E-28) and markers of inflammation such as C-reactive protein (0.026 SD, p= 2.7E-10), Gamma glutamyl transferase (0.035 SD, p= 4.2E-17), glycoprotein acetyls (0.038 SD, p= 3.6E-13) and Alkaline Phosphatase (0.034 SD, p= 3.2E-16) (**Supplemental Data Table 5, Supplemental Figure 10-11**). Cardiac age acceleration was associated with decreased insulin-like growth factor 1 (−0.025 SD, p= 4.6E-09).

Cardiac age acceleration was associated with metabolomic measurements consistent with a healthy diet, such as greater levels of monounsaturated fats (0.027 SD, p= 2.6E-07) (which is likely indicative of increased fat intake generally), a lower degree of unsaturation of the fats measured in the serum (−0.025 SD, p=4.6E-6), and a lower percentage of linoleic acid of total fatty acids (−0.049 SD, p= 1.8E-20) (**Supplemental Data Table 5)**. It was associated with decreased Omega 6 fatty acid percentage of total fatty acids (−0.041 SD, p= 8.7E-15), but not percentage of Omega 3 of total fatty acids (−0.0075 SD, p= 0.15) (**Supplemental Data Table 5)**. It was also associated with increased valine (0.014 SD, p= 6.4E-3), tyrosine (0.021 SD, p= 5.6E-5) and spectrometer corrected alanine (0.017 SD, p= 1.0E-3), but decreased histidine (−0.021 SD, p= 5.5E-5) and glycine (−0.032 SD, p= 6.6E-09), each of which is directionally consistent with associations between serum levels of these amino acids and CVD^36–38^ (**Supplemental Data Table 5)**.

### Association of Cardiac Age Acceleration with Serum Proteins

We tested associations between cardiac age acceleration and serum proteins collected at UK Biobank enrollment. Of the 1,463 proteins tested, cardiac age acceleration was significantly associated with 239 serum proteins at a false discovery rate of 5%, 20 of which were also Bonferroni significant (**Supplemental Figure 12, Bulk Data Table 1**). Some biomarkers of note include natriuretic peptide B (NPPB), NTproBNP, Growth Differentiation Factor 15 (GDF15), Prostasin (PRSS8) and

Asialoglycoprotein Receptor 1 and some protective biomarkers such as Carbonic Anhydrase 14 (CA14), Carbonic Anhydrase 6 (CA6) and Kit Ligand (KITLG). To explore disease patterns related to discovered biomarkers, they were also clustered by disease association, which is explored in more detail in the **Supplemental Results.**

### Disease PheWAS of Cardiac Age Acceleration

To explore the impact of prior disease on cardiac age acceleration, we explored associations between cardiac age acceleration and prevalent disease at the time of imaging (**Supplemental Figures 15, Supplementary Data Table 6, Bulk Data Tables 2)**. Cardiac age acceleration was associated with prevalent cardiovascular specific diseases such as dilated cardiomyopathy (DCM) (Effect size of 0.65, p=2.7E-04), aortic stenosis (0.42, p= 7.2E-07), mitral regurgitation (0.28, p= 2.1E-05), atrial fibrillation (0.29, p= 6.0E-27), and coronary artery disease (0.23, p= 1.4E-35). It was also associated with risk factors such as hypertension (HTN) (0.28, p= 6.9E-227), hyperlipidemia (HLD) (0.12, p= 4.6E-34), and type 2 diabetes (0.27, p= 2.5E-41); diseases that share cardiovascular risk factors such as chronic obstructive pulmonary disease (COPD) (0.17, p= 3.5E-11) and stroke (0.21, p= 1.6E-05); and inflammatory conditions such as systemic lupus erythematosus (SLE) (0.37, p= 1.1E-04) (**Supplemental Data Table 6**).

To explore future disease risk of participants with cardiac age acceleration, we explored associations between cardiac age acceleration and disease incident to imaging (**Supplemental Figure 16, Supplemental Data Table 7, Bulk Data Table 3**). It was most strongly associated with incident hypertrophic cardiomyopathy (hazard ratio of 2.58, p= 6.57E-07. It is also associated with mitral valve disease (1.30, p= 1.03E-07) and aortic valve disease (1.40, p= 1.41E-09) and atrial fibrillation or flutter (1.18, p= 1.60E-06). Greater cardiac age acceleration was also associated with an increased risk of all-cause mortality (1.22, p= 2.16E-07) (**Supplemental Data Table 7)**.

A sensitivity analysis was performed which showed that a Cox model to predict heart failure from cardiac age acceleration incident to MRI including age and sex as covariates was significant (1.08, p=5.6E-6) and as still significant after the addition of LVEF and RVEF as covariates (1.07, p=6.9E-5). Further results exploring the impact of accounting of MRI IDPs on disease prediction can be found in the **Supplemental Results**.

### GWAS of Cardiac Age Acceleration

Next, we conducted analyses testing the relationship between common genetic variants and cardiac age acceleration. Cardiac age acceleration as characterized by our model had an estimated heritability of 25.9%. Given the moderate heritability of cardiac age acceleration, a genome wide association study (GWAS) was conducted in 64,076 participants passing quality control (**Supplemental Figure #3**). 24 independent loci were found to be associated with cardiac age acceleration at P<5e-08 (**Figure 3A**). Of these, 20 loci had not previously been associated with cardiac age acceleration ^7,9^.

**Figure 3:**
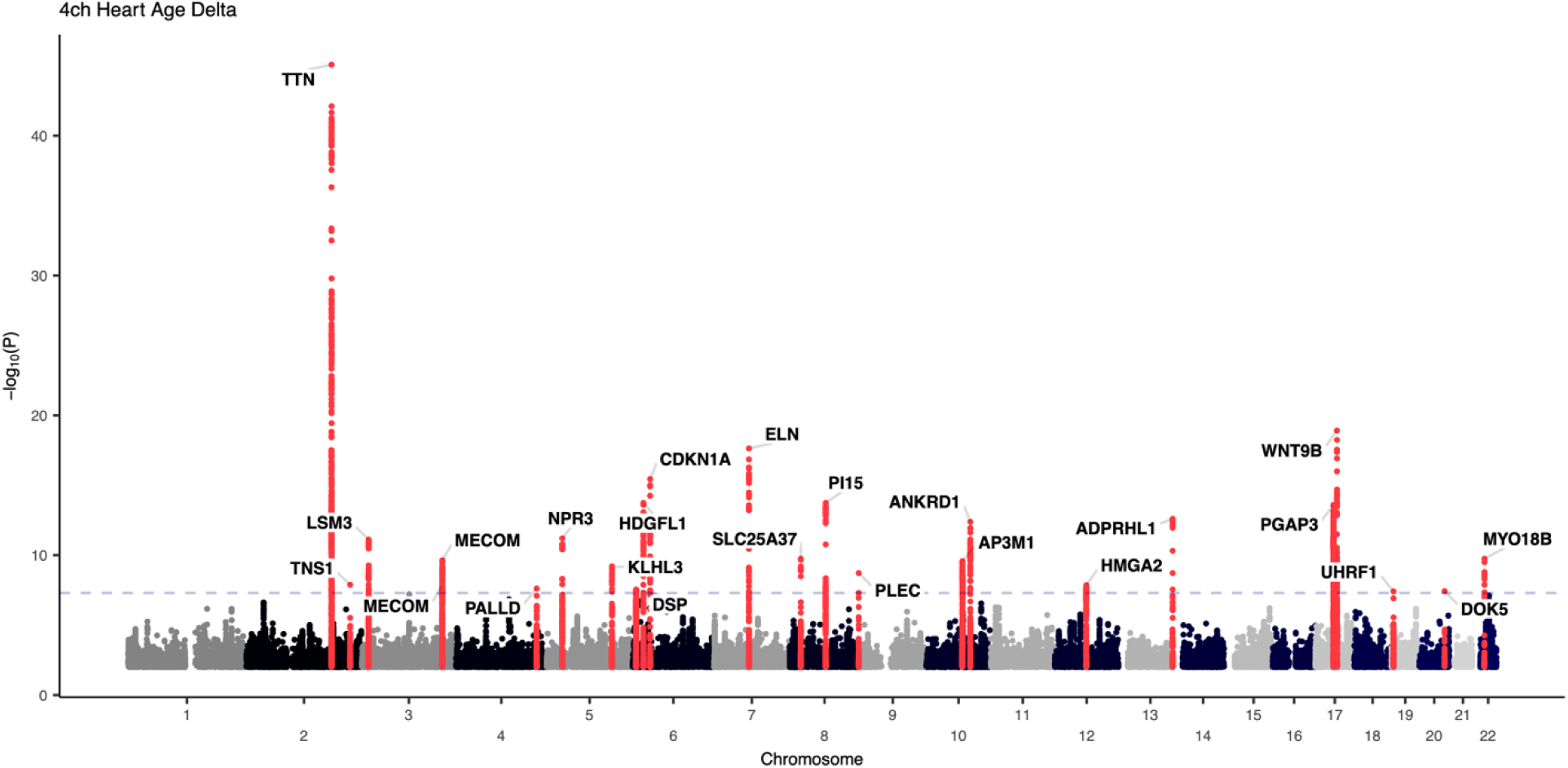
GWAS Manhattan plot for cardiac age acceleration. Manhattan plot of the GWAS of cardiac age acceleration. Lead SNPs are labeled with the nearest gene.

At several loci, the nearest gene to the most strongly associated variant had a known role in contractility or sarcomere function, including *TTN* (titin), *PALLD* (pallidin)^39^, *PLEC* (plectin)^40^, *DSP* (desmoplakin)^41,42^ and *MYO18B* (myosin 18b)^43^. Another subset of genes may play active roles in cardiac development (*MECOM*^44–47^, *ANKRD1*^48^, *ADPRHL1*^49–51^, *WNT9b*^52^). Other loci were near genes relevant to arterial structure and function (*PI15*^53^*, ELN*/elastin)^54^, cellular senescence (*CDKN1A*/p21^55–58^, *HMGA2*^59–61^, *SLC6A6*^62–64^) and proteostasis (*KLHL3*^65,66^*, UHRF1*^67^).

To further explore likely pathways impacted by cardiac age acceleration, MAGMA was used to map GWAS loci to known gene sets. Cardiac age acceleration was associated with gene sets related to valve morphology and function; muscle stretch, cardiac morphogenesis and elasticity. When gene sets were analyzed based on tissue type from GTEx^25^, the most relevant tissue types were heart and arterial tissues (**Figure 4A**). Within cardiac single-cell databases, cardiac age acceleration was most strongly linked to genes expressed in cardiomyocytes and endocardial cells (**Figure 4B**).

**Figure 4:**
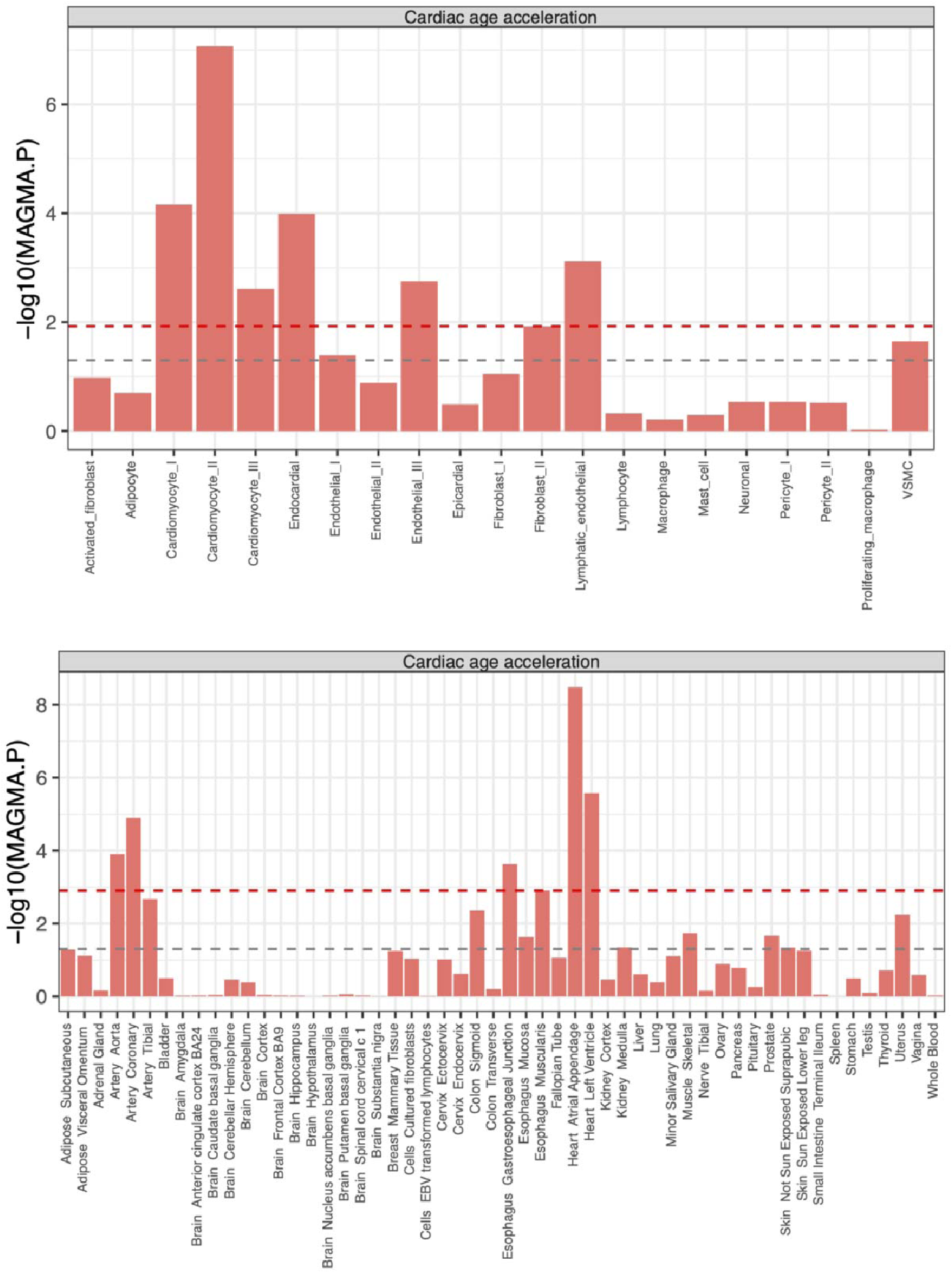
MAGMA gene set associations. *Top:* Associations between MAGMA gene sets and cardiac cell types from Chaffin, *et al*.^26^ *Bottom:* Associations between MAGMA gene sets and tissue expression from GTEx.

### Mendelian Randomization of Cardiac Age Acceleration

Next, we used two-sample Mendelian randomization to explore putatively causal relationships between common exposures like blood pressure, blood counts and lipids (**see Methods for full list**) and cardiac age acceleration. To avoid sample overlap, a GWAS was run on each exposure in a subset of UK Biobank participants without imaging and excluding those within three degrees of kinship of participants with cardiac MRI. We found evidence supporting a causal relationship between greater systolic (0.29, p= 4.7E-14) and diastolic blood pressure (0.31, p= 1.0E-14) and greater age acceleration. Similar observations were made for increased hemoglobin (0.10, p= 6.75E-05), hematocrit (0.11, p= 6.75E-05), which was concordant with the observational findings. Finally, greater cardiac age acceleration was associated with higher levels of lipoprotein(a) (0.02, p= 4.8E-03). Associations with LDL (p=0.01), apolipoprotein B (p=0.02), and apolipoprotein A (p=0.02) were not significant after accounting for multiple testing (**Supplemental Data Table #9, Bulk Data Table 5**).

A proteomic MR was conducted in which cis-genetic instruments for 992 proteins were tested against the outcome of cardiac age acceleration (**Bulk Data Table 4**). Genetically mediated serum levels of 11 proteins were associated with cardiac age acceleration (**Table 2**). Cardiac age acceleration was causally associated with decreased serum levels of proteins associated with cardiac structure and development (R-spondin3^68^, HER-2^69^) but increased levels of proteins was associated with development of atherosclerosis (Furin^70,71^, Latexin^72^). It was also causally associated with serum levels of proteins that may mediate immune impact on cardiovascular disease such as decreased cathepsin C^73–78^ and increased dectin-2^79,80^. The strongest protein in the MR associated with cardiac age deceleration was with peptide methionine sulfoxide reductase, overexpression of which prolongs lifespan in drosophila, but has variable effects on lifespan in mice ^81^. Increased dectin-2 was significantly associated with cardiac age acceleration in both the continuous PheWAS and the MR. HER-2, R-spondin-3 and Furin were associated with increased cardiac age acceleration in the continuous PheWAS, but were causally associated with cardiac age deceleration in the MR.

### Polygenic Predictors of Cardiac Age Acceleration

We sought to explore both the cardiac specificity and disease relevance of the GWAS findings by constructing a polygenic score that estimated cardiac age acceleration. The cardiac age acceleration polygenic predictor was significantly associated with disease outcomes in a held-out set of 395,620 individuals in the UK Biobank unrelated within 3 degrees of kinship to those who underwent MRI (**Supplemental Data Table #10, Bulk Data Table 6**). The polygenic risk predictor created from the cardiac age acceleration GWAS was associated with atrial septal defect (HR 1.14 per SD change in PRS, p= 2.07E-3). It was also associated with atrial fibrillation (HR 1.08, p= 3.02E-41), conduction system disease (HR 1.06, p= 4.69E-16), bradyarrhythmia (HR 1.06, p= 6.81E-13), coronary artery disease (HR 1.05, p= 7.44E-22), heart failure (HR 1.07, p= 2.48E-17), hypertension (HR 1.05, p= 3.33E-30) and hyperlipidemia (HR 1.04, p= 9.86E-12) (**Supplemental Data Table #10**). The polygenic score was also associated with mortality (HR 1.02, p= 5.09E-05) (**Supplemental Data Table #10**).

### Masking Increases Specificity of Cardiac Age Acceleration

To gain more insight into the impact of masking versus differing modeling approaches compared to prior literature, a single fold of the dataset was trained without masking, i.e., permitted to train on non-cardiac structures in addition to cardiac structures. It achieved an MAE of 2.70, a Pearson R of 0.8978 and an R^2^ of 0.81, a value similar to that previously achieved in a modeling approach that incorporated ECG and 2-, 3-, and 4-chamber MRI data without masking (**Supplemental Table #1**).

To test whether our approach of masking out non-cardiac tissue contributed to the phenotype specificity, a sensitivity analysis was performed to evaluate age acceleration from a model trained on unmasked MRIs. While such an approach yielded a higher heritability, the GWAS based on these un-masked measurements yielded only 4 significant loci, and only one locus was shared with the cardiac specific model (near *TTN*) (**Supplemental Figure #18, Supplemental Data Table 8**), further supporting the importance of masking for the discovery of genetic variation relevant to cardiac-specific aging.

## Discussion

Here, we studied cardiac age acceleration—the apparent divergence between the estimated age of the heart and chronological age—in 66,521 UK Biobank participants. By combining an end-to-end deep learning-based age regression model with a segmentation model that excluded non-cardiac structures, we identified cardiac age acceleration to be a heritable phenotype with a high degree of cardiovascular specificity.

The modeling approach was designed to capture dynamic information about cardiac structure and function, while preventing the model from learning from adjacent non-cardiac tissues. The present approach blended the strengths of previous approaches that estimated age acceleration from cardiac-specific measurements with the expressive power of end-to-end deep learning models to facilitate the extraction of age-relevant information from raw pixel intensity and motion data. The information presented to the model can be visualized as a 208×208px image animated over 50 timepoints during the cardiac cycle (2.2 million points). Because we set the pixel intensities for non-cardiac structures to 0 with our initial deep learning model, the data available to the age regression model was limited to heart size, pixel intensities, and the apparent motion of cardiac structures and the blood pool. These constraints deprived the model of useful but non-specific ancillary information and worsened performance for the training task (estimating chronological age). However, sensitivity analyses confirmed that these constraints imbued the model with cardiovascular specificity, compared to a more liberal approach that permitted non-cardiac structures to be seen by the model.

Importantly, the information contained within our cardiac age acceleration measurements could not simply be accounted for by standard measurements of cardiac structure and function: LV strain and wall thickness, aortic measurements, and volumetric measurements from all four cardiac chambers together explained only 10% of the variance in cardiac age acceleration. Still, cardiac age acceleration was associated with adverse cardiac structure and function—and remained predictive of subsequent cardiovascular diagnoses including aortopathy, valvular heart disease, and cardiomyopathies even after accounting for traditional LV measurements. Similarly, a polygenic score for cardiac age acceleration was predictive of adverse cardiovascular consequences but not of many non-cardiovascular age related diseases like back pain, lung cancer or Parkinson’s Disease (**Bulk Data Table 2 and 3**).

Notably, cardiac age acceleration appeared to serve as a readout of cardiovascular fitness and healthy dietary habits. For example, individuals with greater levels of moderate-to-vigorous physical activity as quantified by accelerometry had reduced cardiac age acceleration. While a more complete understanding of the mechanisms by which activity reduces cardiac age acceleration will require future study, we did note that greater genetically mediated circulating levels of the protein product of *RSPO3*, a myokine secreted during skeletal muscle contraction, were inversely associated with cardiac age acceleration in the proteomic Mendelian randomization analysis. Similarly, serum levels of Omega-6, linoleic acid, and increased degree of unsaturation of serum lipids were inversely associated with cardiac age acceleration. These are consistent with dietary recommendations regarding consumption of polyunsaturated fats from the American Heart Association. Adverse behavioral traits, including elevated tobacco and alcohol use to shortened sleep duration, were also linked to cardiac age acceleration. Through these findings, cardiac age acceleration appeared to serve as an embodiment of previously latent information about cardiovascular fitness, which may have implications for future efforts to comprehensively quantify cardiovascular health.

Predisposition to accelerated cardiac aging was found to be heritable and linked to 20 additional genetic loci beyond those that were previously identified. These loci were notable for their cardiovascular specificity: although the deep learning model was forced to estimate age exclusively from cardiac structures, it was nevertheless possible that the main genetic determinants of age acceleration in those structures could have been global senescence and repair-related loci. However, most of the loci appeared to be specific to the cardiovascular system: three loci harbored genes linked to monogenic cardiovascular diseases (*TTN* and *DSP* for cardiomyopathy and *ELN* for aortopathy); four additional loci were near genes with known roles in contractility or sarcomere function (*TNS1*, *PALLD*, *PLEC*, and *MYO18B*); and several other loci were near genes linked to cardiac development (*MECOM*, *ANKRD1*, *ADPRHL1*, *WNT9B*). Tissue-specific gene set analyses underscored the link between cardiac age acceleration and cardiac and proximal arterial tissues (**Figure 3C**); these links were further refined with single-nucleus RNA sequencing analyses that highlighted cardiomyocytes and endothelium as the relevant cell types in the heart (**Figure 3B**) and vascular smooth muscle cells, endothelial cells, and fibroblasts in the aorta. These findings suggest that cardiac age acceleration’s heritable component is predominantly—but not exclusively—linked to variation in its constituent organ systems, rather than to a global aging program.

Still, there were several notable relationships beyond the cardiovascular system. In the GWAS, we identified several loci that may play a more global role in cellular growth and senescence (*CDKN1A*/p21, *HMGA2*, *SLC6A6*/*LSM3*) or proteostasis (*KLHL3*, *UHRF1*) in multiple tissues. Furthermore, inflammatory diseases present before imaging were associated with accelerated cardiac aging (including lupus and rheumatoid arthritis), as were elevated circulating levels of inflammatory markers such as C-reactive protein. Circulating levels of the protein encoded by *CLEC6A*, Dectin-2, and its genetic prediction were both associated with accelerated cardiac aging at P<1E-03. Dectin-2 ligand binding initiates a signal transduction cascade in the pro-inflammatory NFκB/IL-1β pathway. Through these lines of evidence, several sources of inflammation were linked to cardiac age acceleration.

There was also a pattern of association between cardiac age acceleration and adverse changes in brain imaging phenotypes, including findings consistent with diminished cerebral blood flow, gray matter atrophy, increased carotid atherosclerosis, and cerebral small vessel disease. These relationships were not accounted for by risk factors such as blood pressure. These observations add to the growing evidence for a shared heart-brain axis, but because the phenotypes were measured contemporaneously in the same individuals, future efforts will be required to gain insight into causal relationships.

We propose three areas of future interest. First, there is value in continuing to increase the richness of the data that is available to the models, but enforcing cardiac specificity is critical. Second, we expect that significant effort will be required to make the measurements generalize beyond the specific imaging devices on which the models were trained; doing so will be important for generalization and to allow for ongoing characterization of these phenotypes beyond the biobank setting. Third, we expect that some features driving the model estimates of cardiac age acceleration could be parameterized into measurements that can be calculated with more traditional measurement techniques; if so, these features are likely to be more interpretable. Most importantly, some may play a biological role that can be studied independently.

We also note several limitations of our current approach. The calculation of cardiac age acceleration as the difference between calendar age and DL predicted age remains an estimate of structural and functional decline as captured by the DL model. While we believe that it is a stronger proxy than from clinical features alone, it is limited both by what is captured in the MRI and what is extracted by the DL model. Cardiac age acceleration was estimated using 4-chamber long-axis images from cardiovascular MRI in one cohort (UK Biobank) using one device manufacturer. The models were not able to incorporate data from unseen portions of the heart, and are not expected to generalize to other modalities or even to data from other devices without fine-tuning. The cohort was predominantly composed of middle-aged individuals with genetic identities similar to Europeans.

In conclusion, cardiac age acceleration is a heritable, cardiovascular-specific phenotype that embodies information about fitness and disease risk that is not captured in traditional cardiac measurements.

## Supporting information

Supplemental Results

Bulk Data Tables

## Data Availability

GWAS summary statistics for cardiac age acceleration and model weights, along with training and quality control code are available at https://zenodo.org/records/12802434. At publication, final GWAS summary statistics will be added to the GWAS Catalogue, and any code changes will be updated.

## Acknowledgements/Funding

J.N.B. is supported by funding from the Sarnoff Cardiovascular Research Foundation. J.P.P. is supported by NIH K08HL159346. G.H.T. is supported by the National Institutes of Health NHLBI K23HL135274.

