## Supplemental Figures and Tables for "Genetics of Cardiac Aging Implicate Organ-Specific Variation"

Cardiac Age Acceleration Supplemental Tables/Figures

### Supplemental Table #1 Comparison of Model Performance with Selected Literature

| **Citation** | **Input Data** | **Model Style** | **R2** | **MAE** |
| --- | --- | --- | --- | --- |
| Shah, 2023 | Cardiac MRI, 4-chamber view | Imaging-derived phenotypes | **0.49** | **4.2** |
| Raisi-Estabragh, 2022 | Cardiac MRI, 4-chamber view | Radiomics | **0.31** | **5.0** |
| Goallec, 2021 | Cardiac MRI, multiple views | End-to-end, all pixels | **0.78** | **NR** |
| **Present work** | Cardiac MRI, 4-chamber view | End-to-end, masked noncardiac pixels | **0.71** | **3.3** |
|  | Cardiac MRI, 4-chamber view | End-to-end, unmasked | **0.81** | **2.7** |

### Supplemental Table #2 Baseline Characteristics of the Cross Validation Groups

|  |  | **Cross Validation Group** | | | | | | | | | |
| --- | --- | --- | --- | --- | --- | --- | --- | --- | --- | --- | --- |
|  | **Total**  **(N=61691)** | **G0**  **(N=6120)** | **G1**  **(N=6208)** | **G2**  **(N=6224)** | **G3**  **(N=6215)** | **G4**  **(N=6265)** | **G5**  **(N=6193)** | **G6**  **(N=6111)** | **G7**  **(N=6092)** | **G8**  **(N=6103)** | **G9**  **(N=6160)** |
| **Sex** |  |  |  |  |  |  |  |  |  |  |  |
| Female | 31889 (52 %) | 3131 (51 %) | 3212 (52 %) | 3198 (51 %) | 3242 (52 %) | 3211 (51 %) | 3207 (52 %) | 3165 (52 %) | 3169 (52 %) | 3141 (51 %) | 3213 (52 %) |
| Male | 29802 (48 %) | 2989 (49 %) | 2996 (48 %) | 3026 (49 %) | 2973 (48 %) | 3054 (49 %) | 2986 (48 %) | 2946 (48 %) | 2923 (48 %) | 2962 (49 %) | 2947 (48 %) |
| **Age at time of MRI** |  |  |  |  |  |  |  |  |  |  |  |
| Mean (SD) | 66 (± 7.7) | 65 (± 7.7) | 66 (± 7.8) | 66 (± 7.8) | 66 (± 7.7) | 66 (± 7.7) | 66 (± 7.7) | 66 (± 7.7) | 66 (± 7.8) | 66 (± 7.7) | 66 (± 7.8) |
| **BMI** |  |  |  |  |  |  |  |  |  |  |  |
| Mean (SD) | 27 (± 4.5) | 27 (± 4.5) | 27 (± 4.5) | 26 (± 4.5) | 27 (± 4.4) | 27 (± 4.5) | 27 (± 4.6) | 27 (± 4.4) | 27 (± 4.4) | 27 (± 4.4) | 27 (± 4.4) |
| Missing | 253 (0.4%) | 22 (0.4%) | 29 (0.5%) | 29 (0.5%) | 22 (0.4%) | 21 (0.3%) | 21 (0.3%) | 28 (0.5%) | 26 (0.4%) | 29 (0.5%) | 26 (0.4%) |
| **Systolic Blood Pressure mmHg)** |  |  |  |  |  |  |  |  |  |  |  |
| Mean (SD) | 140 (± 19) | 140 (± 20) | 140 (± 19) | 140 (± 19) | 140 (± 19) | 140 (± 19) | 140 (± 19) | 140 (± 19) | 140 (± 20) | 140 (± 19) | 140 (± 19) |
| Missing | 7666 (12.4%) | 769 (12.6%) | 788 (12.7%) | 782 (12.6%) | 752 (12.1%) | 795 (12.7%) | 761 (12.3%) | 757 (12.4%) | 777 (12.8%) | 752 (12.3%) | 733 (11.9%) |
| **Diastolic Blood Pressure (mmHg)** |  |  |  |  |  |  |  |  |  |  |  |
| Mean (SD) | 79 (± 10) | 79 (± 10) | 79 (± 10) | 79 (± 10) | 79 (± 10) | 79 (± 10) | 79 (± 10) | 79 (± 10) | 79 (± 10) | 79 (± 10) | 79 (± 10) |
| Missing | 7666 (12.4%) | 769 (12.6%) | 788 (12.7%) | 782 (12.6%) | 752 (12.1%) | 795 (12.7%) | 761 (12.3%) | 757 (12.4%) | 777 (12.8%) | 752 (12.3%) | 733 (11.9%) |

### Supplemental Figure #1 Model Calibration and Regression Dilution Bias Adjustment

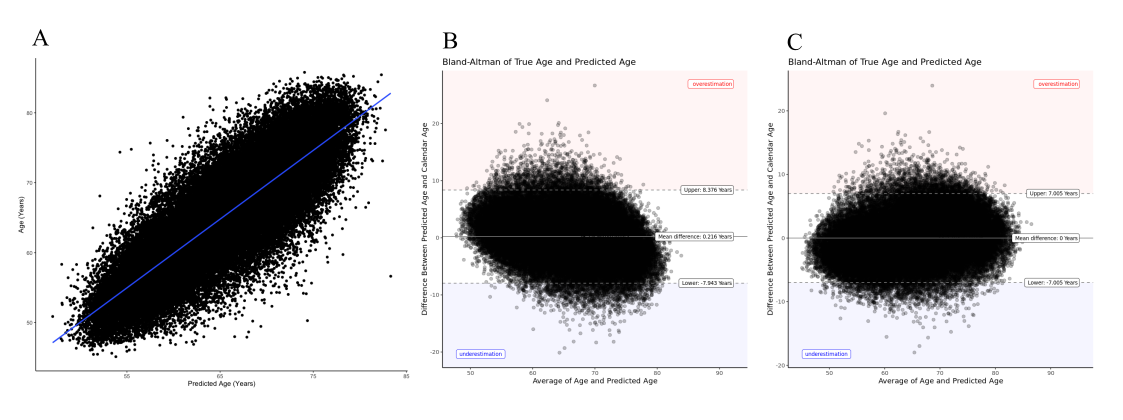
Cardiac Age Regression Model Calibration: (A) Scatterplot of predicated age versus calendar age. Before adjustment for regression dilution bias, age is overestimated in younger participants and underestimated in older participants (B). After adjustment for regression dilution bias, that trend is mitigated (C)

### Supplemental Table #3 Comparison of Each Validation Fold on Universal Holdout Set

| **Metric** | **G0** | **G1** | **G2** | **G3** | **G4** | **G5** | **G6** | **G7** | **G8** | **G9** |
| --- | --- | --- | --- | --- | --- | --- | --- | --- | --- | --- |
| MAE | 3.3100 | 3.3339 | 3.3414 | 3.3377 | 3.3611 | 3.3807 | 3.1575 | 3.3092 | 3.2411 | 3.3289 |
| Pearson R | 0.8410 | 0.8349 | 0.8327 | 0.8419 | 0.8343 | 0.8320 | 0.8512 | 0.8382 | 0.8434 | 0.8290 |
| RMSE | 4.1858 | 4.2719 | 4.2965 | 4.2636 | 4.2826 | 4.2730 | 4.0620 | 4.2124 | 4.1655 | 4.3225 |
| R-Squared | 0.7073 | 0.6971 | 0.6934 | 0.7088 | 0.6961 | 0.6922 | 0.7245 | 0.7026 | 0.7113 | 0.6872 |
| Performance of the Full Model on the Universal Validation Set | | | | | | | | | | |

### Supplemental Figure #2 Alignment Quality Control

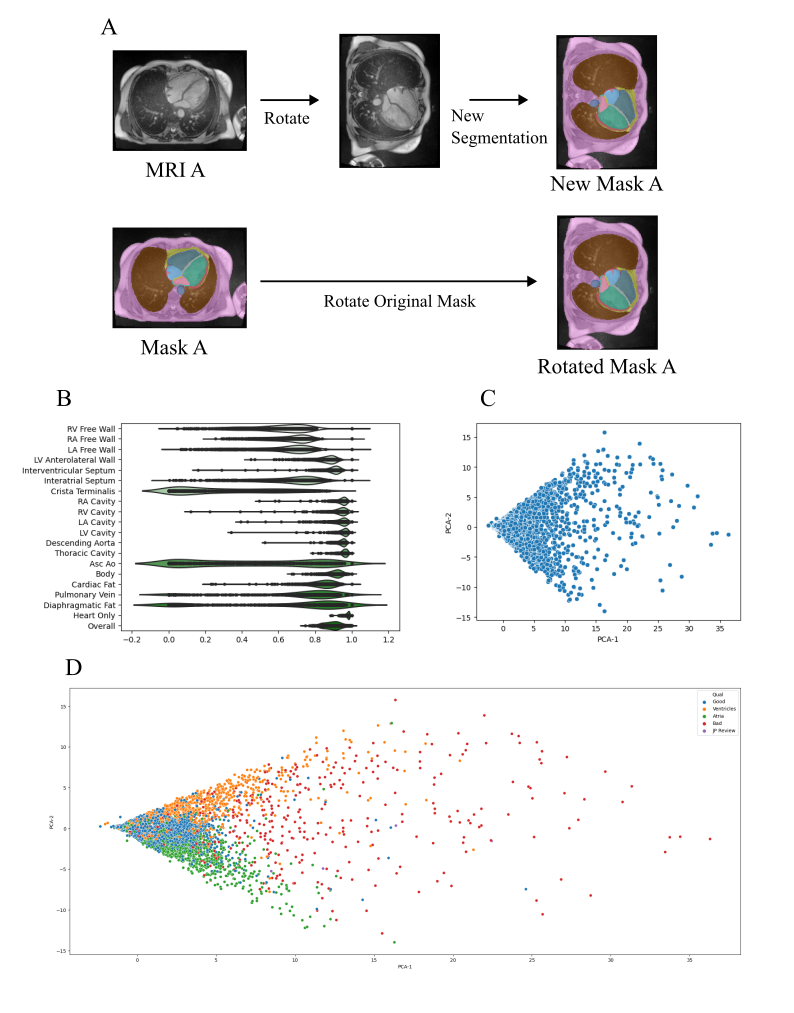

Alignment Quality Control: (A) Display of the alignment workflow. An MRI and its mask are rotated by some number of degrees (90 degrees here) and the rotated MRI is used to generate a new mask. The new mask is compared with the original rotated mask and per region DICE scores are calculated. (B) Displays the distributions of DICE scores in each region. Then PCA of the regions is performed, with the first two dimensions shown in (C). All MRIs with components outside the two standard deviations from of the mean of the first two components were manually visualized and given a quality score. High scores in PCA-1 tended to have poor alignment in all regions (D). MRIs with high values of PCA-2 tended to have misalignment of the ventricles while lower PCA-2 values were associated with atrial misalignment (D).

### Supplemental Table #4: List of Cardiac MRI Image Derived Phenotypes

**Image Derived Phenotypes**

f_LVenddiastolicvolume

f_LVendsystolicvolume

f_LVstrokevolume

f_LVejectionfraction

f_LVcardiacoutput

f_LVmyocardialmass

f_RVenddiastolicvolume

f_RVendsystolicvolume

f_RVstrokevolume

f_RVejectionfraction

f_LAmaximumvolume

f_LAminimumvolume

f_LAstrokevolume

f_LAejectionfraction

f_RAmaximumvolume

f_RAminimumvolume

f_RAstrokevolume

f_RAejectionfraction

f_Ascendingaortamaximumarea

f_Ascendingaortaminimumarea

f_Ascendingaortadistensibility

f_Descendingaortamaximumarea

f_Descendingaortaminimumarea

f_Descendingaortadistensibility

f_LVmeanmyocardialwallthicknessAHA1

f_LVmeanmyocardialwallthicknessAHA2

f_LVmeanmyocardialwallthicknessAHA3

f_LVmeanmyocardialwallthicknessAHA4

f_LVmeanmyocardialwallthicknessAHA5

f_LVmeanmyocardialwallthicknessAHA6

f_LVmeanmyocardialwallthicknessAHA7

f_LVmeanmyocardialwallthicknessAHA8

f_LVmeanmyocardialwallthicknessAHA9

f_LVmeanmyocardialwallthicknessAHA10

f_LVmeanmyocardialwallthicknessAHA11

f_LVmeanmyocardialwallthicknessAHA12

f_LVmeanmyocardialwallthicknessAHA13

f_LVmeanmyocardialwallthicknessAHA14

f_LVmeanmyocardialwallthicknessAHA15

f_LVmeanmyocardialwallthicknessAHA16

f_LVmeanmyocardialwallthicknessglobal

f_LVcircumferentialstrainAHA1

f_LVcircumferentialstrainAHA2

f_LVcircumferentialstrainAHA3

f_LVcircumferentialstrainAHA4

f_LVcircumferentialstrainAHA5

f_LVcircumferentialstrainAHA6

f_LVcircumferentialstrainAHA7

f_LVcircumferentialstrainAHA8

f_LVcircumferentialstrainAHA9

f_LVcircumferentialstrainAHA10

f_LVcircumferentialstrainAHA11

f_LVcircumferentialstrainAHA12

f_LVcircumferentialstrainAHA13

f_LVcircumferentialstrainAHA14

f_LVcircumferentialstrainAHA15

f_LVcircumferentialstrainAHA16

f_LVcircumferentialstrainglobal

f_LVradialstrainAHA1

f_LVradialstrainAHA2

f_LVradialstrainAHA3

f_LVradialstrainAHA4

f_LVradialstrainAHA5

f_LVradialstrainAHA6

f_LVradialstrainAHA7

f_LVradialstrainAHA8

f_LVradialstrainAHA9

f_LVradialstrainAHA10

f_LVradialstrainAHA11

f_LVradialstrainAHA12

f_LVradialstrainAHA13

f_LVradialstrainAHA14

f_LVradialstrainAHA15

f_LVradialstrainAHA16

f_LVradialstrainglobal

f_LVlongitudinalstrainSegment1

f_LVlongitudinalstrainSegment2

f_LVlongitudinalstrainSegment3

f_LVlongitudinalstrainSegment4

f_LVlongitudinalstrainSegment5

f_LVlongitudinalstrainSegment6

f_LVlongitudinalstrainglobal

f_Averageheartrate

f_Cardiacoutput

f_Cardiacindex

### Supplemental Table #5 Model Performance of Each Validation Fold on its own Validation Set

| **Metric** | **G0** | **G1** | **G2** | **G3** | **G4** | **G5** | **G6** | **G7** | **G8** | **G9** | **Overall** |
| --- | --- | --- | --- | --- | --- | --- | --- | --- | --- | --- | --- |
| MAE | 3.2666 | 3.2753 | 3.2689 | 3.3686 | 3.2832 | 3.2521 | 3.2626 | 3.3721 | 3.2612 | 3.2629 | **3.2873** |
| Pearson R | 0.8434 | 0.8501 | 0.8464 | 0.8404 | 0.8421 | 0.8471 | 0.8452 | 0.8415 | 0.8432 | 0.8443 | **0.8431** |
| RMSE | 4.1457 | 4.1414 | 4.1515 | 4.2735 | 4.1514 | 4.0990 | 4.1492 | 4.2511 | 4.1602 | 4.1623 | **4.1688** |
| R-Squared | 0.7113 | 0.7227 | 0.7164 | 0.7063 | 0.7091 | 0.7176 | 0.7144 | 0.7081 | 0.7110 | 0.7128 | **0.7108** |
| Performance of the Full Model on each test group | | | | | | | | | | | |

### Supplemental Figure #3 CONSORT Diagram

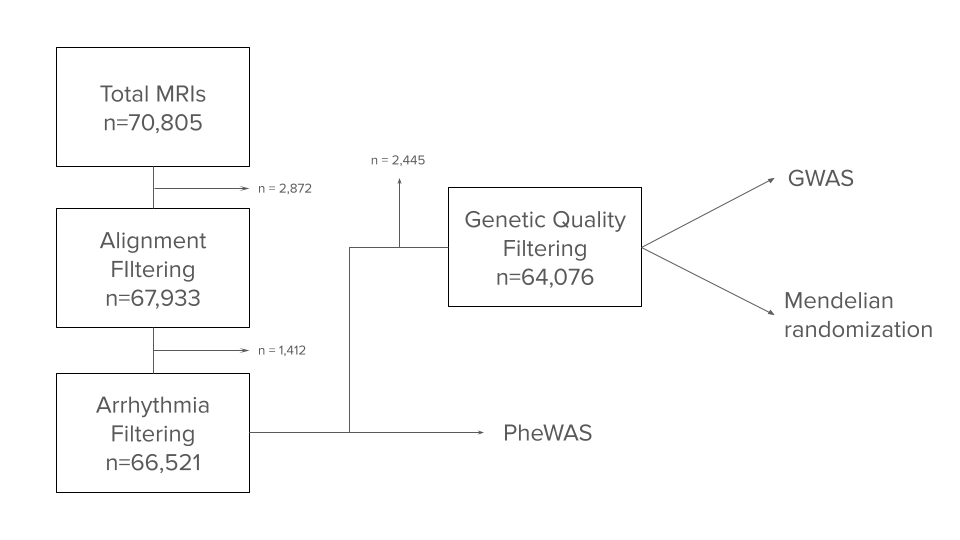

### Supplemental Figure #4 Body Size and Composition Continuous PheWAS

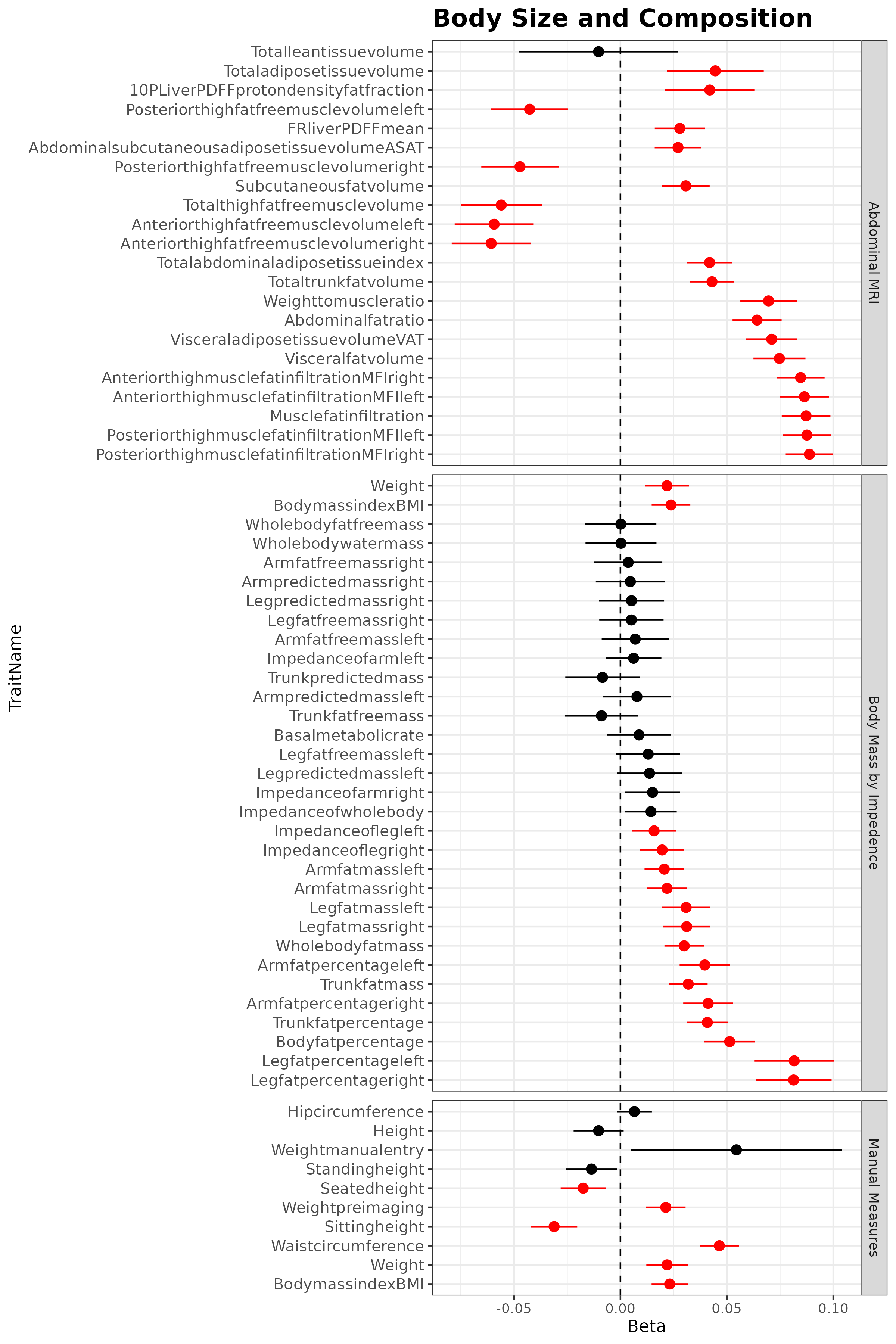

### Supplemental Figure #5: T1 Brain MRI Continuous PheWAS

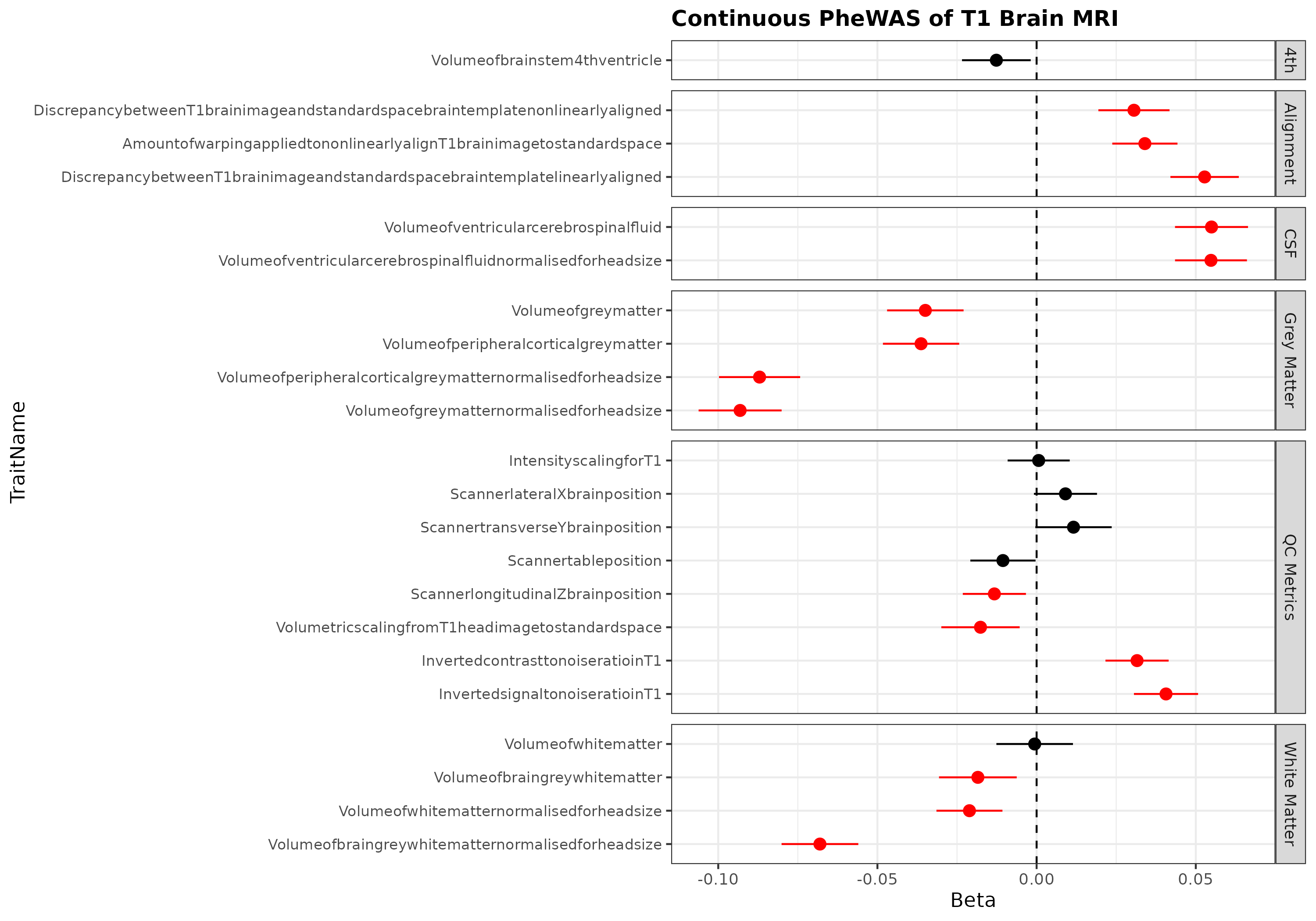

### Supplemental Figure #6: T2/FLAIR Brain MRI Continuous PheWAS

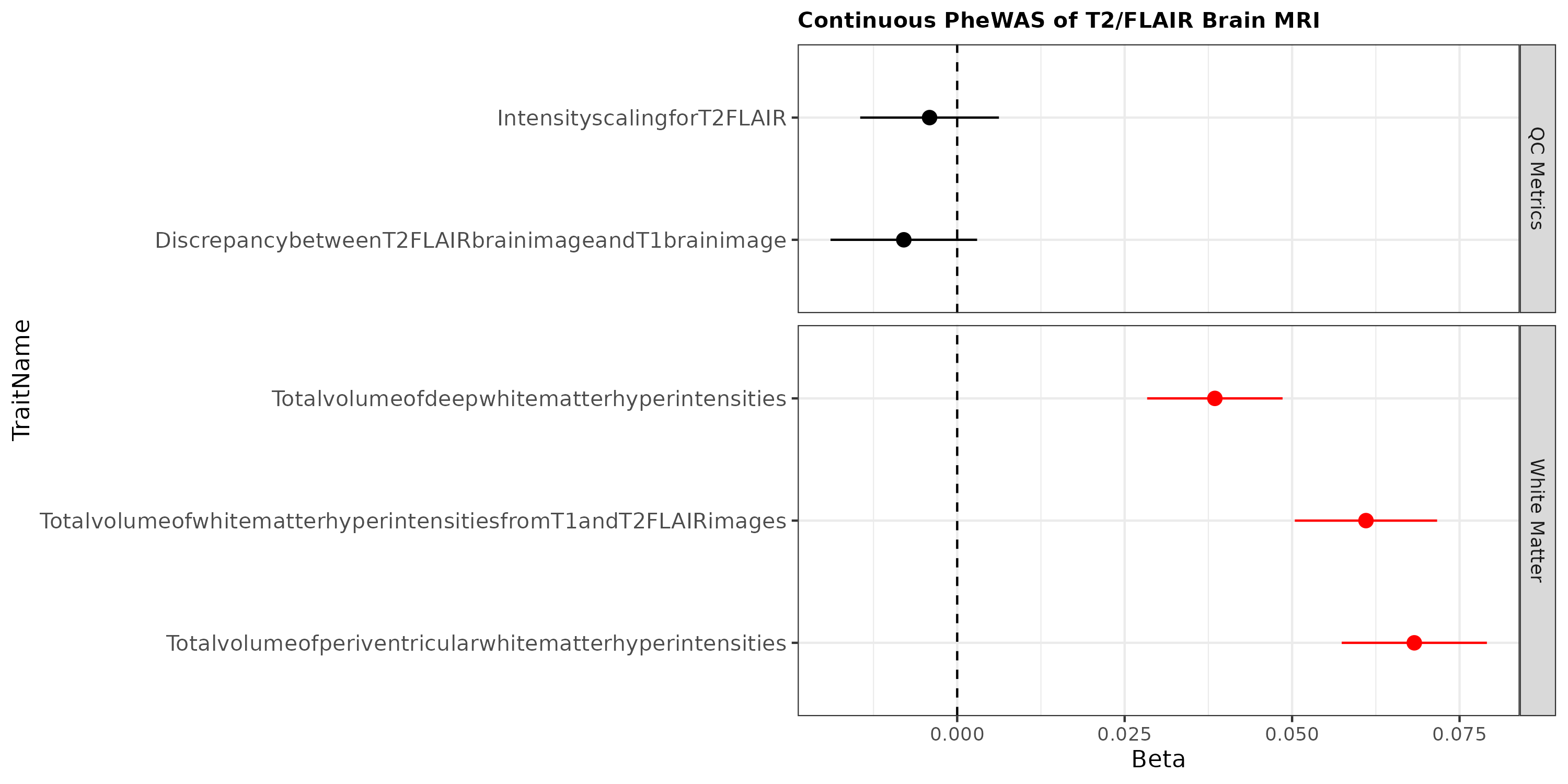

### Supplemental Figure #7: Arterial Spin Continuous PheWAS

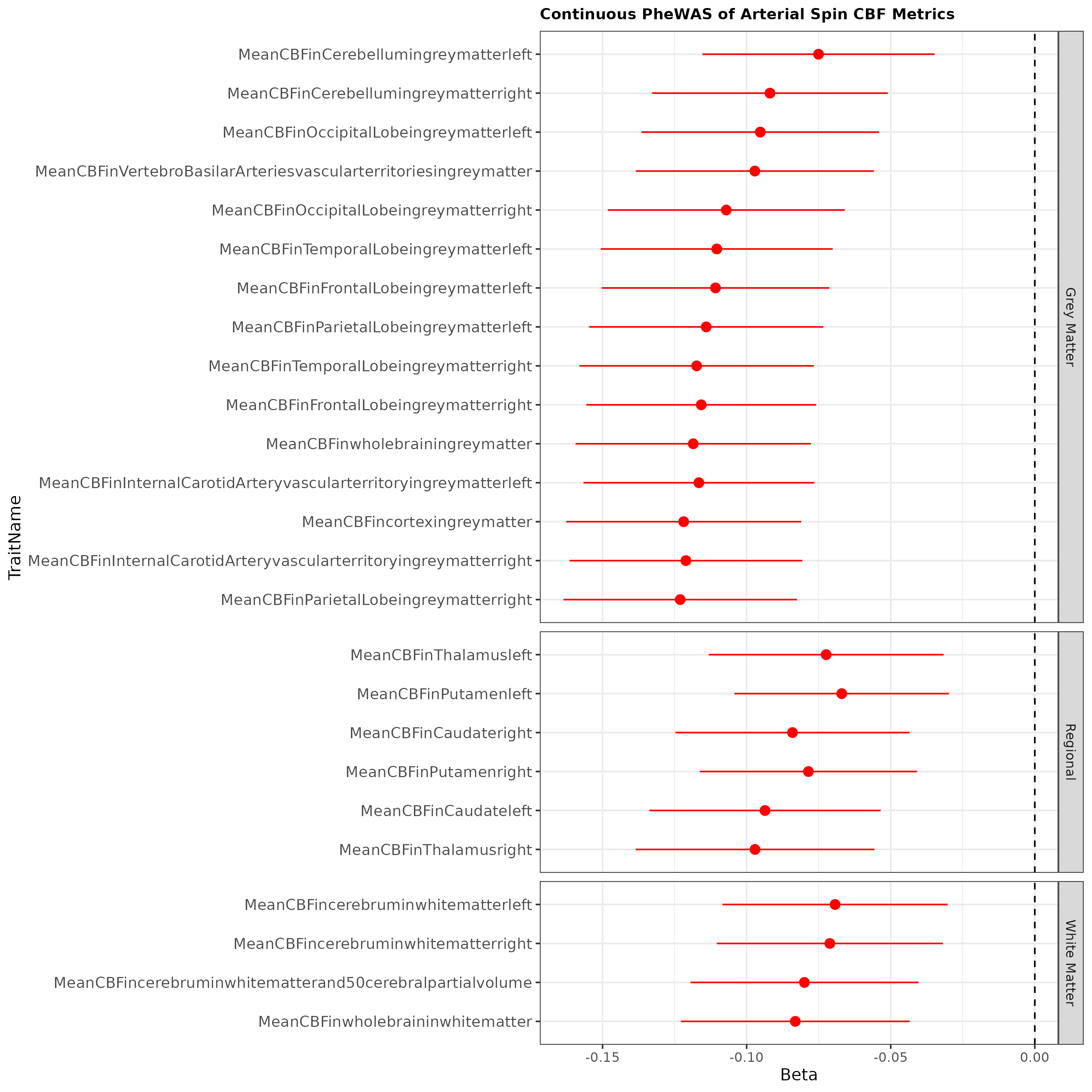

### Supplemental Figure #8: T2*/SWI Brain MRI Continuous PheWAS

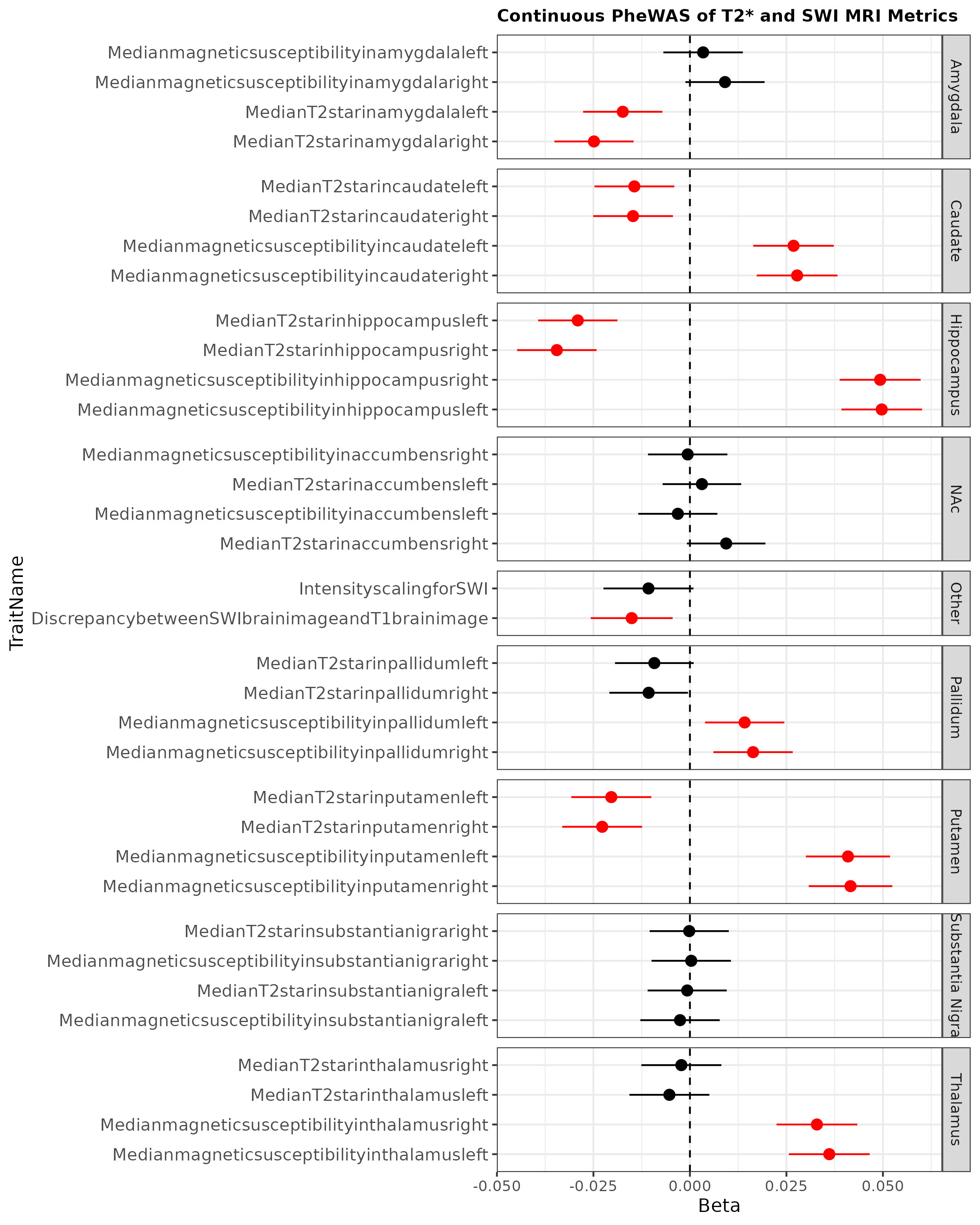

### Supplemental Figure #9: Carotid Ultrasound Continuous PheWAS

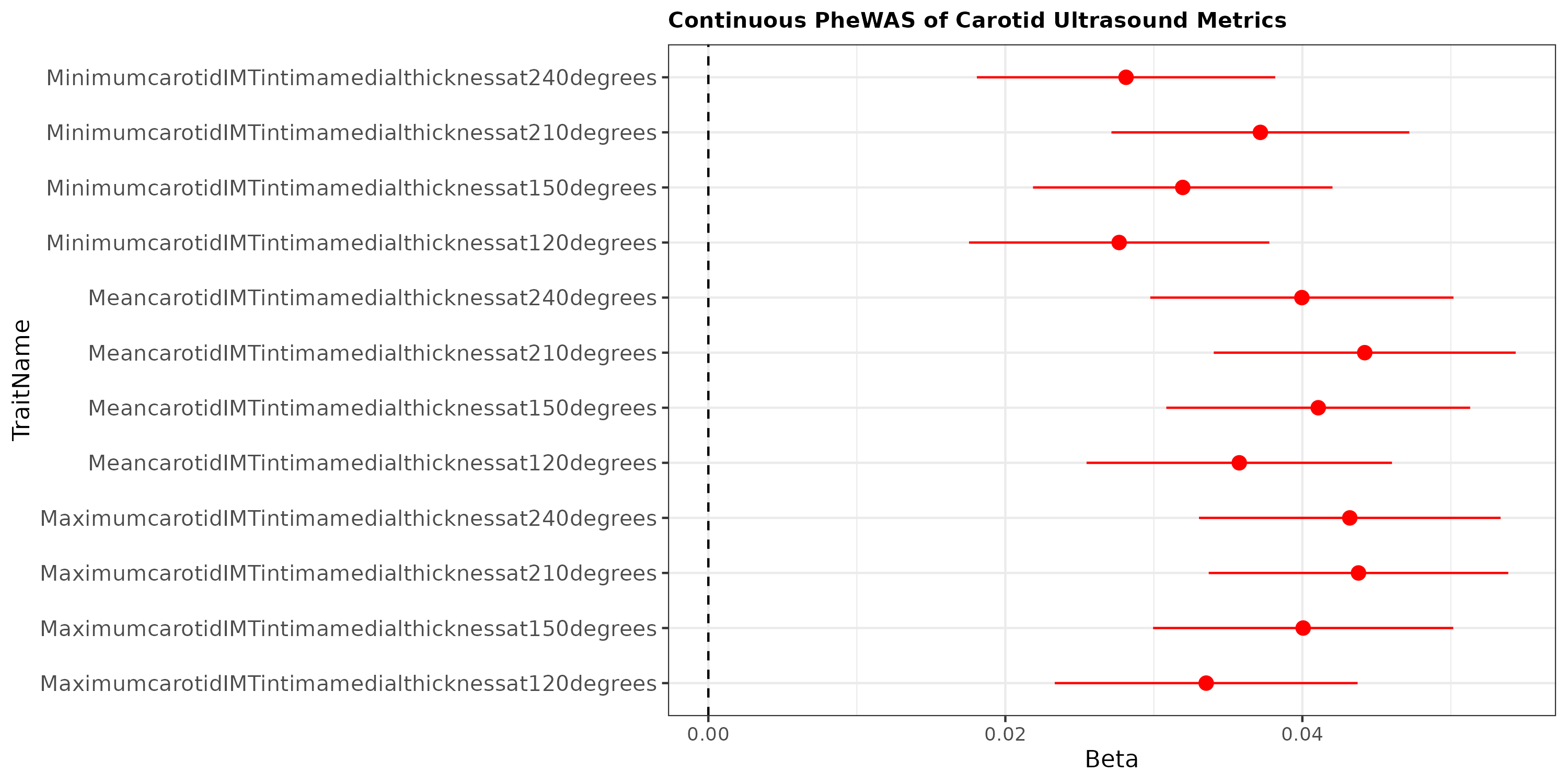

### Supplemental Figure #10: CBC Continuous PheWAS

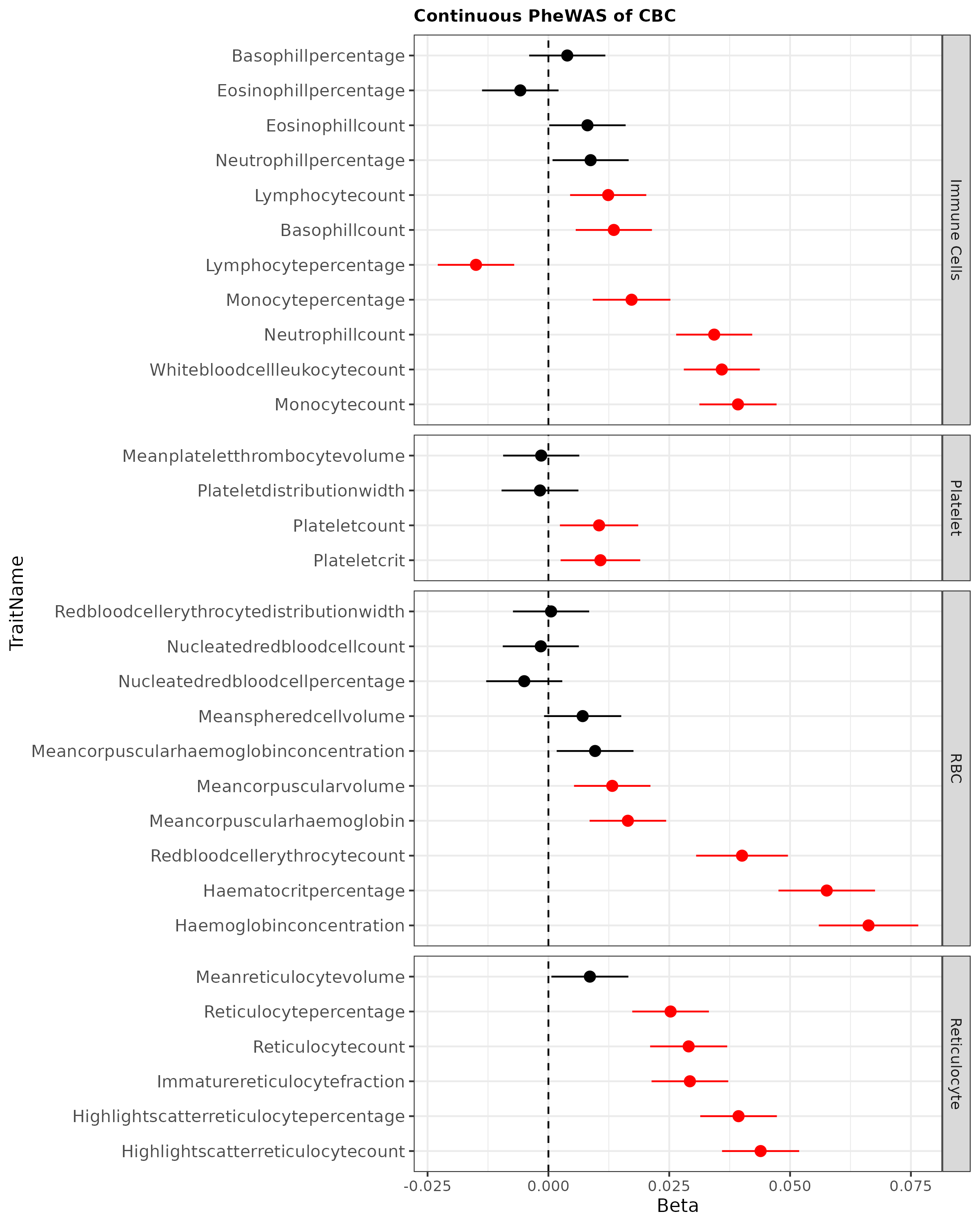

### Supplemental Figure #11 Serum Chemistry Continuous PheWAS

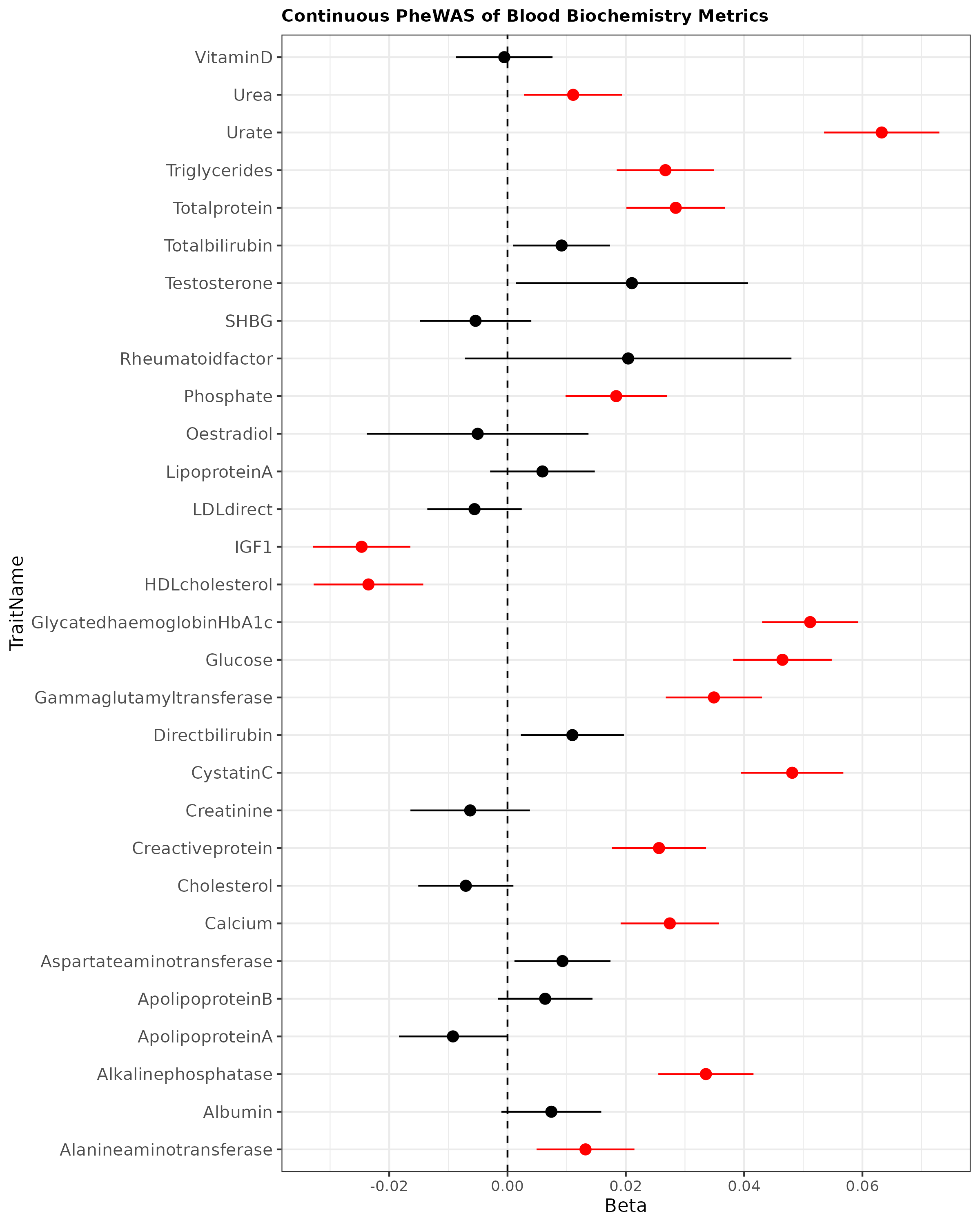

### Supplemental Figure #12 Volcano Plot of Cardiac Age Acceleration Serum Protein Associations

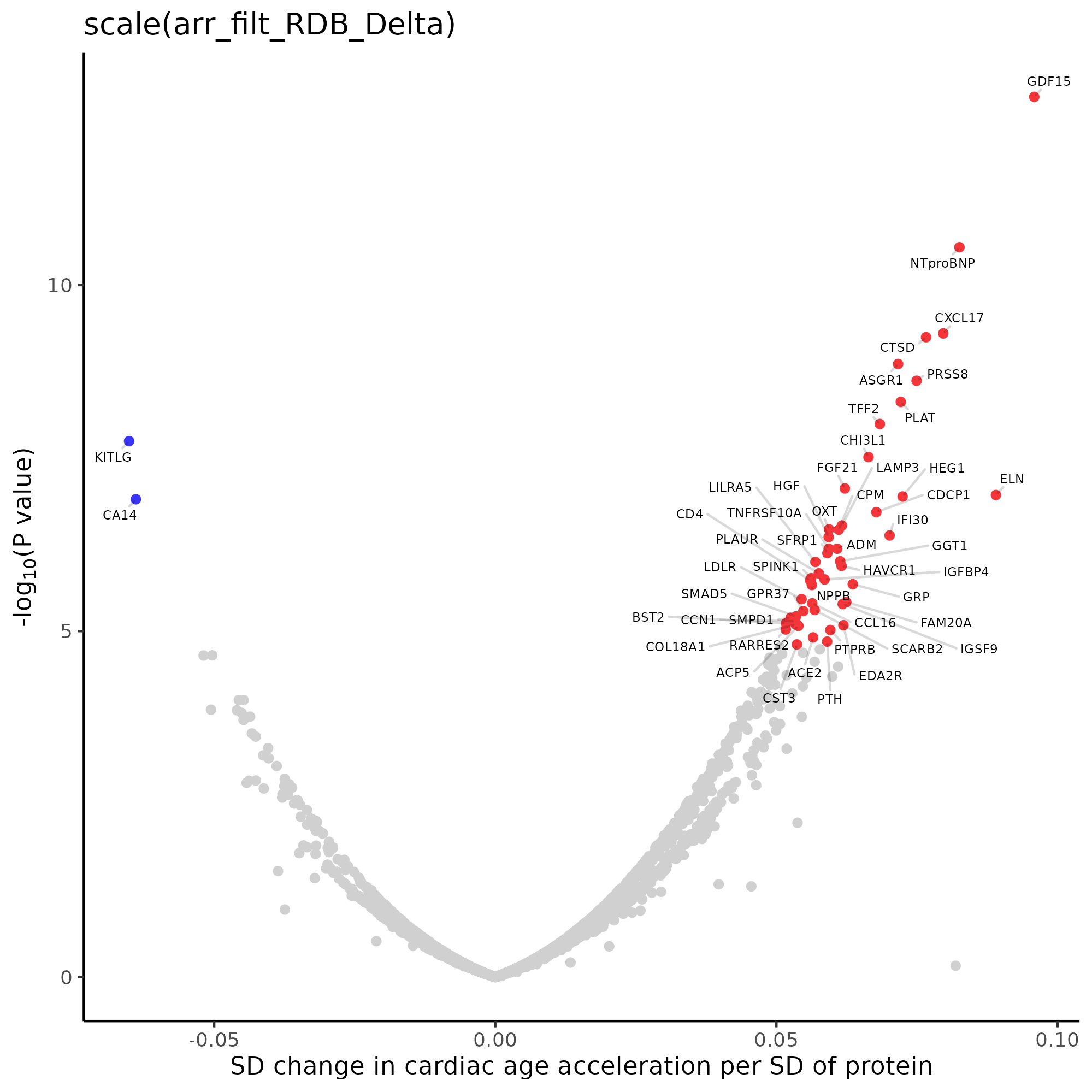

### Supplemental Figure #13 Serum Biomarkers Clustered by Prevalent Disease Associations

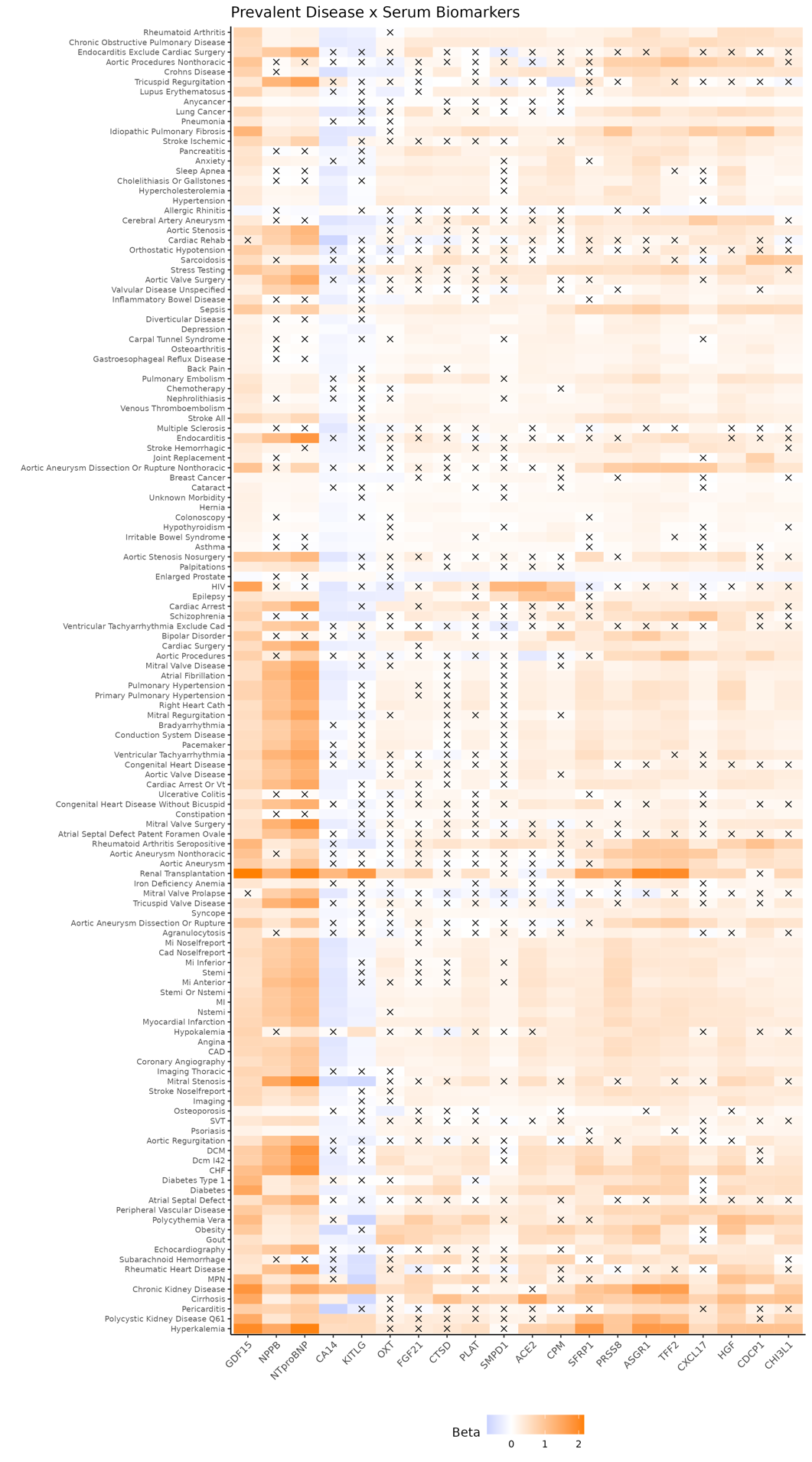

### Supplemental Figure #14 Serum Biomarkers Clustered by Incident Disease Associations

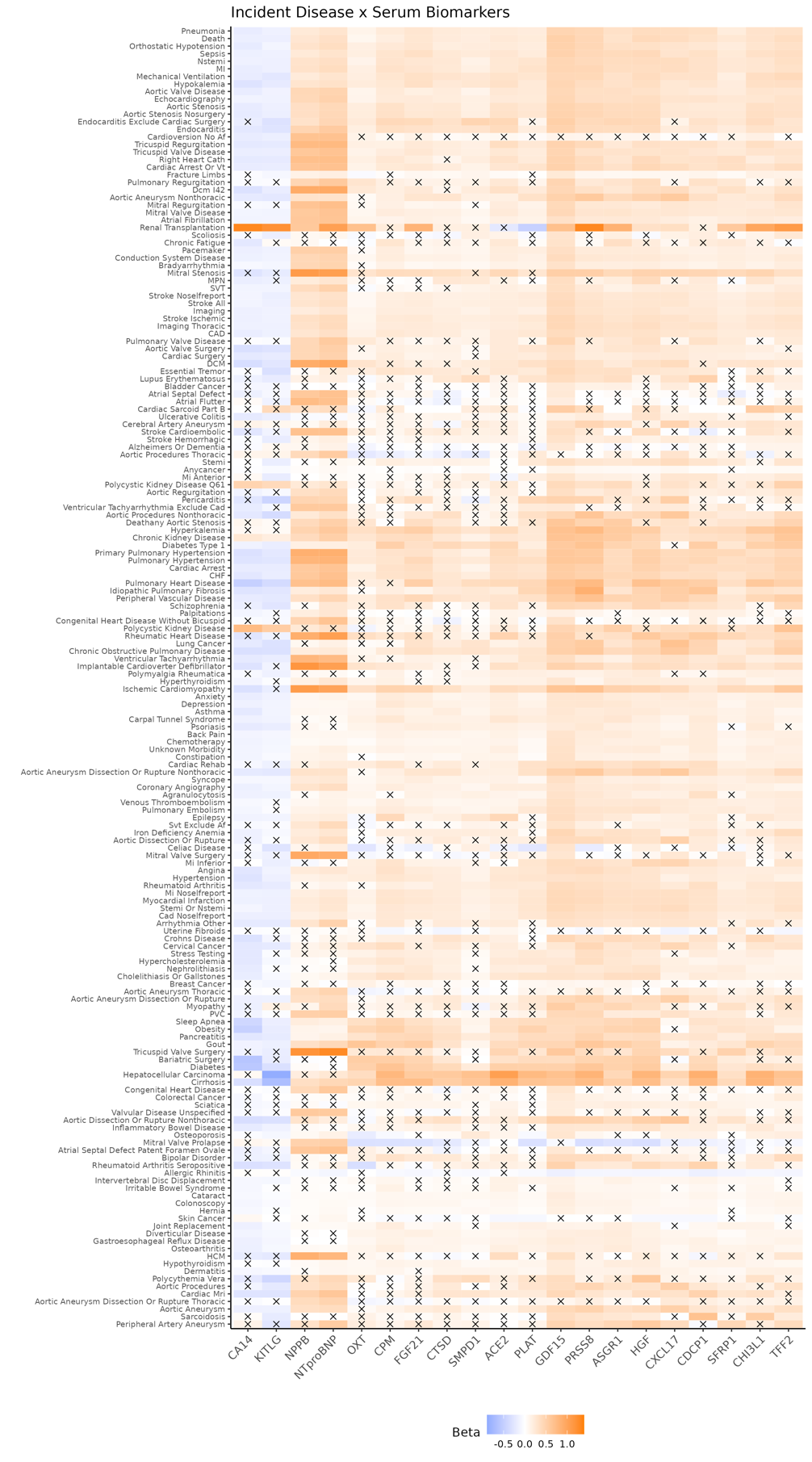

### Supplemental Figure #15 Significant Associations in the Prevalent Disease PheWAS

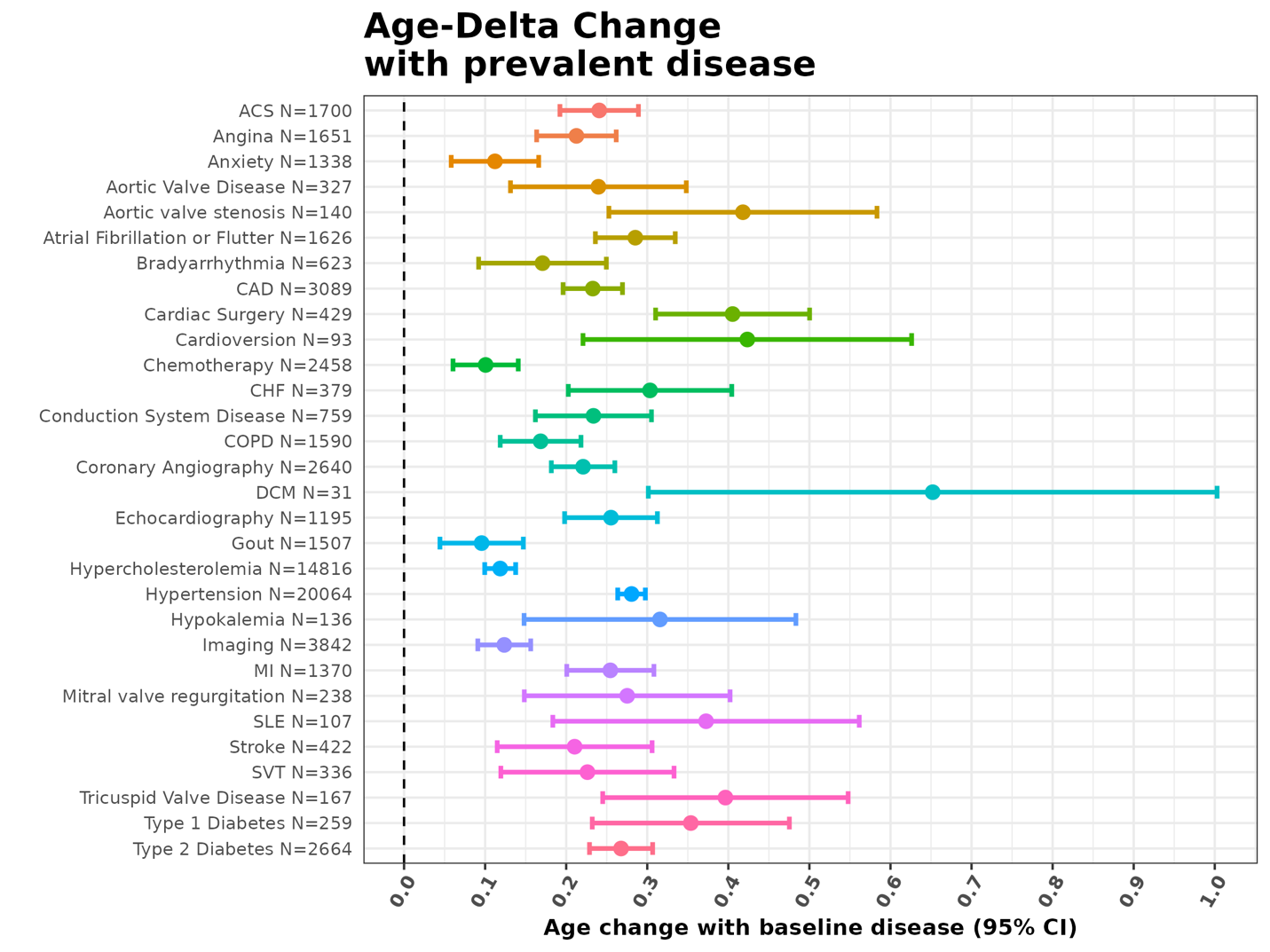

### Supplemental Figure #16 Significant Associations in the Incident Disease PheWAS

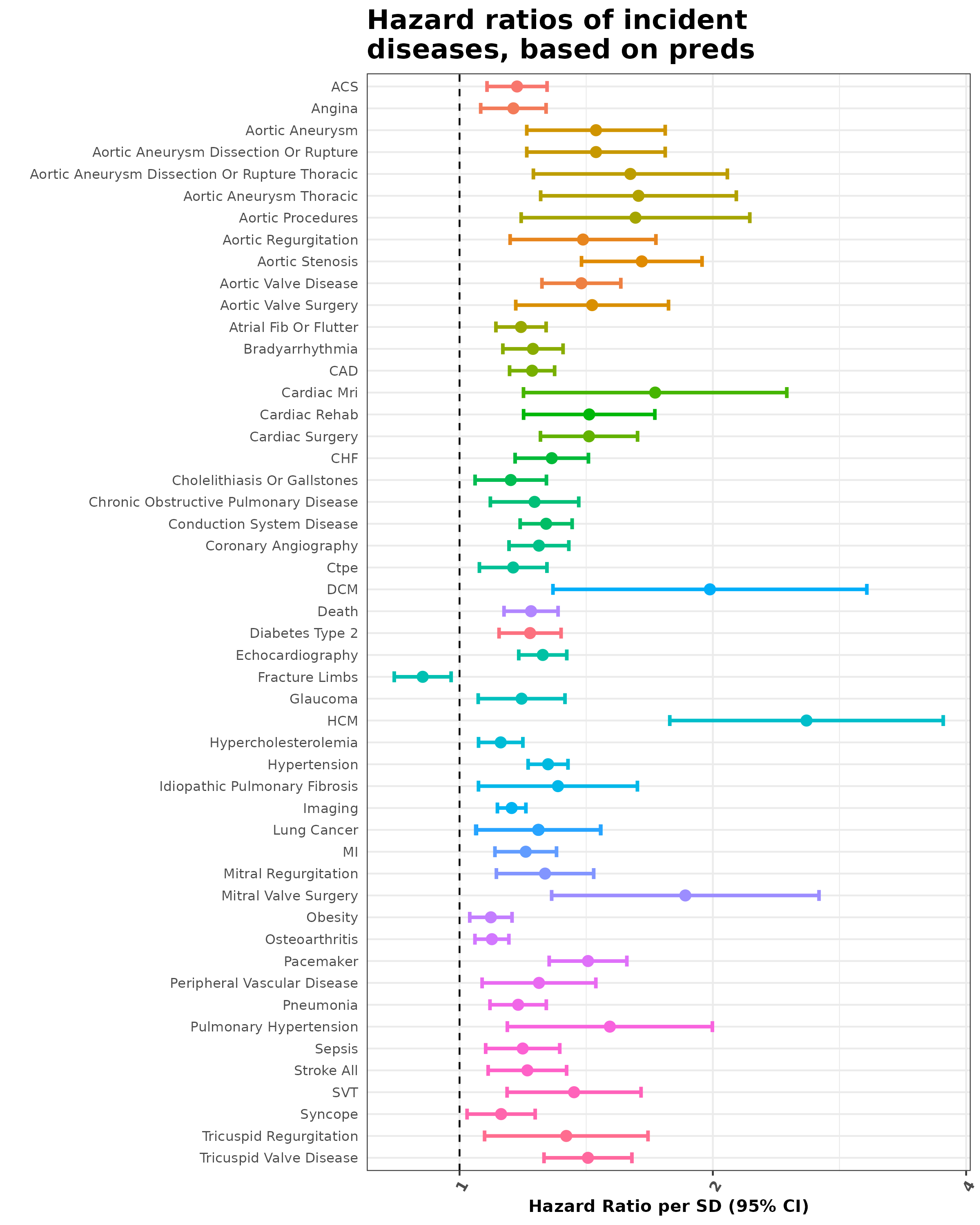

### Supplemental Figure #17 Prevalent PheWAS Compared to Prevalent PheWAS Adjusted for CMR Image Derived Phenotypes

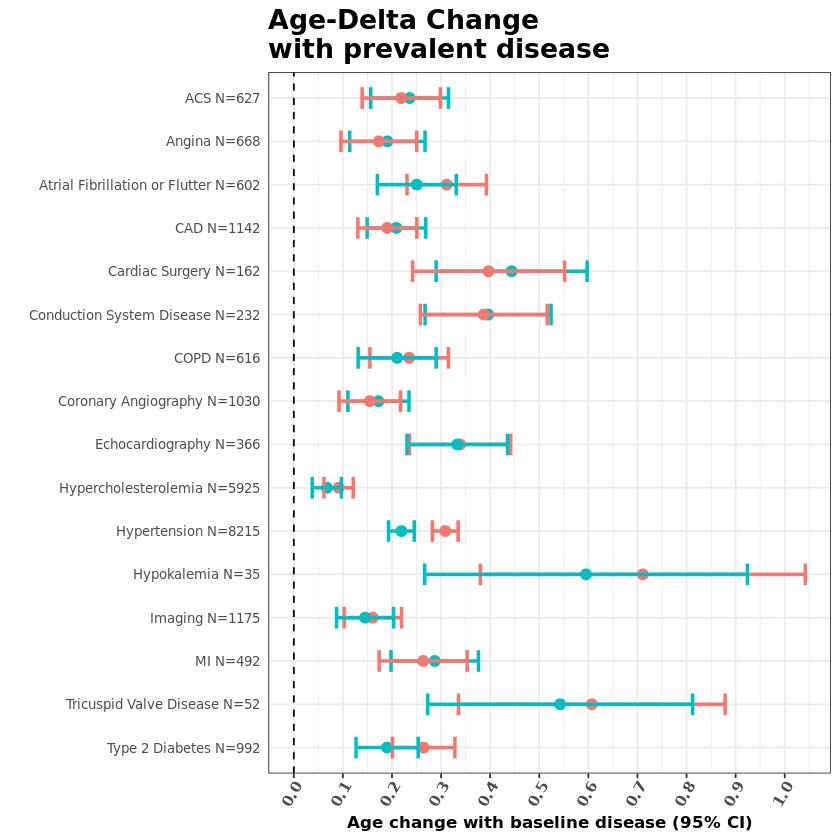

After adjusting for CMR image derived phenotypes (Red), the prevalent disease PheWAS is similar to the original PheWAS when not adjusted for image derived phenotypes (Blue).

### Supplemental Figure #18 GWAS Sensitivity Analysis Mask vs. No Mask

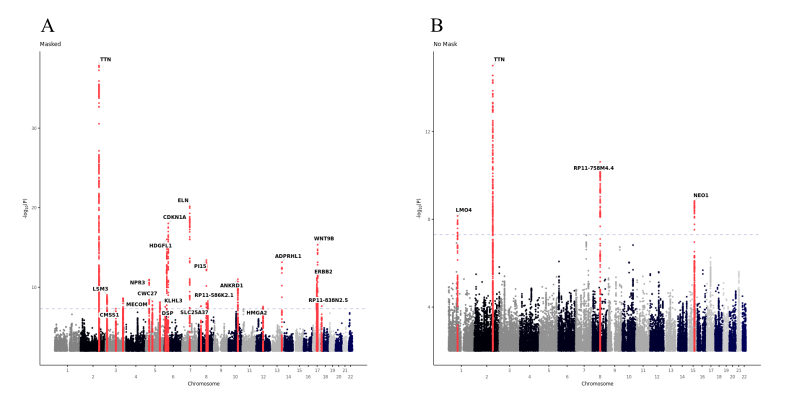

Masking out non-cardiac tissue detects more loci than non-specific models: Manhattan plots visualizing a GWAS from a single fold of the cardiac tissue specific (A) and unmasked MRI (B) models.
