## Supplemental Methods for "Genetics of Cardiac Aging Implicate Organ-Specific Variation"

**MRI Acquisition**

MRI acquisition has been previously described by the UK Biobank^1^. In brief, images were acquired on clinical wide-bore 1.5 Tesla scanners (MAGNETOM Aera, Syngo Platform VD13A, Siemens Healthcare, Erlangen, Germany) using balanced steady-state free precession with typical parameters.

**Genome Wide Association Study Input Quality Control**

In addition to the quality control exclusions noted above, participants were excluded from the GWAS if they lacked genetic data, if the genetic data had a missingness >2%, if there was sex chromosome aneuploidy, or if the genetic data had excessive relatedness to others in the cohort.

**Olink proteomic profiling**

The UK Biobank provided data characterizing 1,463 proteins in the plasma proteome of 54,306 participants using Olink technology^2^. Olink uses antibodies bound to oligomers which—when brought into proximity due to antibody binding—hybridize and extend, allowing the protein to be quantitated via sequencing^3^.

**Trait Exposure GWAS**

Mendelian randomization was performed for the following other traits: systolic blood pressure, diastolic blood pressure, hemoglobin concentration, hematocrit percentage, lipoprotein a, urea, red blood cell erythrocyte count, white blood cell leukocyte count, HDL cholesterol, monocyte count, urate, highlight scatter reticulocyte count, IGF-1, SHBG, cystatin-c, apolipoprotein a, reticulocyte count, platelet crit, triglycerides, nucleated red blood cell percentage, eosinophill count, platelet count, calcium, gamma glutamyltransferase, reticulocyte percentage, BMI, highlight scatter reticulocyte percentage, glucose, aspartate aminotransferase, platelet distribution width, smoker, estradiol, basophill count, c-reactive protein, mean corpuscular haemoglobin concentration, albumin, monocyte percentage, testosterone, total protein, vitamin d, eosinophill percentage, ldl direct, mean sphered cell volume, neutrophill count, alanine aminotransferase, pulse, apolipoprotein b, lymphocyte count, neutrophill percentage, weight, cholesterol, lymphocyte percentage, phosphate, alkaline phosphatase, direct bilirubin, mean reticulocyte volume, total bilirubin, basophill percentage, glycated haemoglobinhb A1c, mean corpuscular haemoglobin, creatinine, immature reticulocyte fraction, red blood cell erythrocyte distribution width, rheumatoid factor, mean corpuscular volume.
