## Supplemental Results for "Genetics of Cardiac Aging Implicate Organ-Specific Variation"

*Impact of Regression Dilution Bias*

Prior to correction for regression dilution bias, the cardiac age delta was associated with calendar age (Beta= -0.28 [approximately -2.8 years per decade], p=1e-5065, **Supplemental Figure 1B**). Cardiac age acceleration, the regression dilution bias-adjusted cardiac age delta, was still associated with calendar age but with a much smaller effect size (Beta= -0.007 (approximately -25.5 days per decade), p=2.1e-5, **Supplemental Figure 1C**). It did not differ between men and women (p=0.91, **Figure 1**). Cardiac age acceleration can thus be defined as the portion of the difference between chronological and model predicted age that cannot be estimated by chronological age alone.

*Cardiac age acceleration is not strongly mediated by image derived phenotypes*

To explore what portion of the disease associations could be attributed to cardiac MRI image derived phenotypes, a linear model to detect prevalent disease which included imaging features and cardiac age acceleration was created. Disease associations were similar after accounting for image derived phenotypes (**Supplemental Figure #17**).

*Clustering of Serum Biomakers by Disease PheWAS*

To understand the relevant conditions that might link those 20 protein markers to cardiac age acceleration, a Cox model was fit to link each individual protein to incident disease occurring after UK Biobank enrollment (**Supplemental Figure 13, Supplemental Figure 14)**. The resulting associations were clustered to explore similar groups of serum biomarkers. A set of biomarkers including natriuretic peptide B (NPPB) and NTproBNP were strongly associated with incident congestive heart failure, HCM and multiple forms of valve disease. Another cluster including Growth Differentiation Factor 15 (GDF15), Prostasin (PRSS8) and Asialoglycoprotein Receptor 1 (ASGR1) was associated with incident cirrhosis, chronic kidney disease and inflammatory disease like gout and rheumatoid arthritis. Finally, another cluster including Carbonic Anhydrase 14 (CA14), Carbonic Anhydrase 6 (CA6) and Kit Ligand (KITLG) was associated with protective effects in many diseases.
